# Transmission dynamics of the COVID-19 pandemic across the emerging variants in mainland China: a hypergraph-based spatiotemporal modeling study

**DOI:** 10.64898/2026.04.16.26351004

**Authors:** Yi Wang, Dong Wang, Yiu-Chung Lau, Zhanwei Du, Benjamin J. Cowling, Yi Zhao, Sheikh Taslim Ali

## Abstract

Mainland China experienced multiple waves of COVID-19 pandemic during 2020-2022, driven by emerging variants and changes in public health and social measures (PHSMs). We developed a hypergraph-based Susceptible–Vaccinated–Exposed–Infectious–Recovered–Susceptible (SVEIRS) model to reconstruct epidemic dynamics across 31 provinces, capturing transmission heterogeneity associated with clustered contacts. We assessed key characteristics of transmission at national and provincial levels during four outbreak periods: initial, localized pre-delta, Delta, and widespread Omicron, which accounted for 96.7% of all infections. We found significant diversity in transmission contributions across cluster sizes, with a small fraction of larger clusters responsible for a disproportionate share of infections. Counterfactual analyses showed that reducing cluster-size heterogeneity, while holding overall exposure constant, could have lowered national infections by 11.70–30.79%, with the largest effects during Omicron period. Ascertainment rates increased over time but remained spatially heterogeneous with a range: (14.40, 71.93)%. Population susceptibility declined following mass vaccination (to 42.49% in Aug 2021, nationally) and rebounded (to 89.89% in Nov 2022) due to waning immunity with variations across the provinces. Effective reproduction numbers displayed marked temporal and spatial variability, with higher estimates during Omicron. Overall, these results highlight critical role of group contact heterogeneity in shaping epidemic dynamics.

## Introduction

The COVID-19 pandemic first emerged in Hubei, China in January 2020 and rapidly spread across all provinces in mainland China. In response, China implemented a “zero-COVID” strategy until November 2022, aiming to eliminate domestic transmission [1,2]. During this period, multiple epidemic waves were driven by emerging variants, evolving interventions, and heterogeneous population immunity. While early containment effectively suppressed nationwide outbreaks, later waves exhibited pronounced provincial heterogeneity in timing, intensity, and control outcomes. Accurately reconstructing transmission at the provincial level is therefore essential for understanding China’s complex epidemic dynamics and the mechanisms underlying spatially heterogeneous spread.

The transmission of COVID-19 was strongly overdispersed, with a small fraction of infected individuals accounting for a disproportionate share of secondary infections, while most cases generate few or no onward transmissions [3–5]. For instance, in Hong Kong, approximately 19% of cases were responsible for 80% of local transmission [3]. Empirical contact-tracing studies consistently showed that large outbreaks are frequently driven by cluster transmission occurring in specific social and environmental contexts, such as retail settings, close-contact indoor social venues, and care homes [6]. Individuals traveling via public transportation or working and living in shared workplaces were repeatedly identified as high-risk populations for Severe Acute Respiratory Syndrome Coronavirus 2 (SARS-CoV-2) transmission [7]. Such clusters involve repeated contacts within shared environments and prolonged exposure, where infection risk depends on the cumulative duration and intensity of interactions rather than merely the number of infectious individuals. For example, digital contact tracing data showed that transmissions occur disproportionately during prolonged exposures (≥1h) rather than brief contacts, implying cumulative exposure effects on transmission probability [8], and household studies consistently found high secondary attack rates in settings with sustained daily contact [9]. As a result, transmission risk increased nonlinearly with contact intensity and duration, making linear dependence on the number of infectious individuals an inadequate approximation of within-cluster transmission dynamics. These observations indicated that overdispersed transmission is not merely a consequence of individual-level variability, but emerges from heterogeneous contact structures and group interaction processes shaped by shared environments and repeated exposure. Despite extensive efforts to quantify transmission overdispersion and its epidemiological consequences, most existing modeling frameworks treat overdispersion as a phenomenological property, typically encoded through a dispersion parameter or stochastic individual-level heterogeneity. While such approaches can reproduce observed variability in case counts, they do not explicitly represent how structured contact environments and cumulative exposure within clusters generate highly uneven transmission outcomes.

Importantly, cluster transmission could not, in general, be faithfully reduced to independent pairwise interactions [10]. When infection risk depends nonlinearly on collective exposure intensity and duration, transmission dynamics are governed by group composition and interaction structure rather than by the sum of dyadic contacts. Models based solely on pairwise interactions therefore lacked the structural resolution required to represent how clustered contact environments give rise to the unevenness of transmission.

Recent studies in social contagion modeling highlighted the role of group interactions, in which transmission occurs through hypergraph-based rather than dyadic contacts, and showed that nonlinear infection processes can fundamentally alter spreading dynamics, giving rise to discontinuous transitions and bistable regimes [10–12]. These phenomena arose from collective exposure mechanisms that are directly relevant to cluster-driven infectious disease transmission. However, existing group contagion frameworks largely remained theoretical and have rarely been integrated with epidemiological incidence data. Consequently, their potential for mechanistically reconstructing real-world transmission heterogeneity was not yet fully realized. This gap motivated the development of data-driven epidemic models that explicitly incorporate group contact structures.

Building on these insights, we developed a hypergraph-based network transmission framework to investigate heterogeneous cluster contact patterns across 31 provinces in mainland China. By integrating nonlinear, cluster transmission dynamics into an SVEIRS compartmental model, we provided a mechanistic reconstruction of COVID-19 spread in mainland China from January 2020 to November 2022. This framework enabled us to identify regional and temporal differences in cluster transmission and to better understand the emergence of superspreading events, thereby offering a basis for context-specific prevention and control strategies.

## Materials and Methods

### Sources of Data

Province-level COVID-19 surveillance data for mainland China were obtained from two authoritative sources: the Johns Hopkins University Center for Systems Science and Engineering (JHU CSSE) [13] and the World Health Organization (WHO) dataset [14]. The JHU CSSE compiles daily reports of confirmed cases on province level, defined as symptomatic individuals with positive PCR test results, based on publicly released data from the National Health Commission (NHC) of the People’s Republic of China and provincial health authorities. In contrast, the WHO dataset aggregated daily nationwide totals reported by the NHC, which encompassed both confirmed symptomatic and laboratory confirmed asymptomatic infections. To obtain the total number of reported infections (including asymptomatic cases) on the province level, we proportionally redistributed the national-level WHO data across provinces based on the spatial distribution of confirmed cases from the JHU CSSE dataset. In this study, we defined the analysis period as spanning from 1 January 2020 to 10 November 2022, the date on which the COVID-19 response was officially optimized to the “20 Measures” policy, which included shortened quarantine periods for international arrivals and a reduced number of personnel subject to quarantine or health monitoring. We considered four distinct periods (waves) of COVID-19 pandemic in mainland China based on the emergence of dominant variants and shifts in transmission dynamics. The *initial outbreak* period occurred before 31 March 2020, followed by the *pre-Delta* period from 1 April 2020 to 11 April 2021. The *Delta* period spanned 12 April 2021 to 21 November 2021, and the *Omicron* period covered 22 November 2021 to August 2022.

Data on COVID-19 vaccine administration in mainland China were obtained from Our World in Data (OWD) [15]. This dataset included the nationwide total number of individuals who received at least one dose, those who completed the initial vaccination protocol, and the total number of booster doses administered, as reported by the NHC of People’s Republic of China (Table S1). Booster doses referred to vaccinations administered in addition to those required by the original vaccination schedule (two doses). We derived the daily counts of first doses, second doses, and booster doses from the difference of these three datasets. Similarly, we allocated vaccine doses to each province proportionally, based on the population distribution across provinces. The vaccination effectiveness is set as a piecewise function (Figure S1). Details on data preprocessing procedures are provided in the Supplementary Material Section 1.2.

Province-level stringency indexes enacted by governments in mainland China were accessed through the Oxford Covid-19 Government Response Tracker (OxCGRT) [16]. The stringent index of province *i* at day *t* reported a number between 0 to 100 that reflected the level of the government’s response, including containment and closure policies, economic policies, health system policies, and vaccination policies. Higher values of the OxCGRT index were assumed to correspond to stricter measures.

The human mobility data between provinces was obtained from the Baidu Migration platform [17]. The source of Baidu migration data was the massive location service data of Baidu Map Open Platform, which came from all software that chooses to use Baidu’s location-based API service. The dataset provided two key indicators. The first is the migration scale index of each province, which in this study referred to the in-migration index, reflecting the scale of inflow to each province. Following a previous study that estimated the scaling coefficient of the migration index [18], we converted the migration index into an approximate count of migrants by applying a coefficient of *δ* = 3.09 × 10^5^. The second is the proportion of inflow to each province from other provinces, indicating the distribution of migrants’ origins. The inter-provincial mobility flows in mainland China on 13 February 2020, the peak of the first pandemic wave, are illustrated in Figure S2. Additional details on data preprocessing procedures are provided in the Supplementary Material Section 1.3.

### Hypergraph-based Compartment Model

We built a metapopulation hypergraph-based SVEIRS model to simulate the transmission dynamics of COVID-19 in mainland China at the province level by considering movement between provinces (Table S2 and Figure S3). The structure of SVEIRS adopted in this study is illustrated in Supplementary Material Section 2.1.

To consider heterogeneous and clustered contact patterns for the infectious transmission, we incorporated a hypergraph, a mathematical framework into the force of infection specifically that captured interactions within clusters, beyond simple pairwise contacts. In the hypergraph-based contact network, individuals are connected through clusters, and susceptible individuals can become infected via adjacent infectors. In a *m*-size cluster containing *k* infected individuals, the force of infection to a susceptible individual was as follows [12],

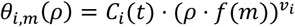

Here, *C*_*i*_ (*t*) is the nonlinear infection proportionality parameter for province *i* at day *t*, 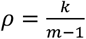 is the density of infected individuals in the *m*-size cluster, and *f*(*m*) = *m* − 1 is the typical number of contacts in environments. *v*_*i*_ is the nonlinear infection kernel for province *i*, related to the heterogeneous exposure periods distribution and the decreased velocity of virus doses [12]. Illustrative examples of the force of infection to a susceptible in the *m*-size cluster are shown in Figure S4.

Assuming the number of infectious individuals in a *m*-size household cluster follows a binomial distribution with parameter 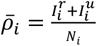 denoting the density of infectious individuals in the entire population, the expected force of infection to a susceptible individual in a *m*-size cluster 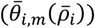 was calculated as,

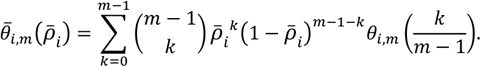

Then, based on the cluster size distribution 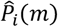 we derived the hypergraph-based force of infection as follows,

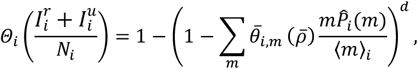

where *d* is the average degree of an individual, ⟨*m*⟩_*i*_ is the mean cluster size of province *i*, 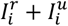 is the sum of reported infections and unreported infections of province *i*, and *N*_*i*_ is the population of province *i*. A contact survey conducted in Chinese provinces outside Hubei after the 2020 lockdown showed that the average degree of individual *d* = 1.19 [19], additional details on degree data were provided in the Supplementary Material Section 1.4. In our model, the cluster size distribution 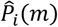 was approximately derived from the household size distribution data from census records in province *i* of mainland China [20] (Figure S5).

We introduced an ascertainment rate parameter (*α*_*i*_(*t*)) in our model, defined as the proportion of total infections that are detected and reported, to account for the underreporting of SARS-CoV-2 infections due to limited testing, asymptomatic cases, or surveillance limitations. It was assumed to be a monotonically increasing function to reflect the improved capability to capture COVID-19 cases over the course of the pandemic. We also accounted for reporting delays in infection by convoluting the modelled incidence with a reporting delay distribution, which was assumed to follow a gamma distribution with a mean of *T*_*d*_ = 6 days and a standard deviation of 19.46 days [21]. The details of the hypergraph-based compartment model and the convolution for the reporting delay were provided in Supplementary Material Section 2.6.

### Effect of Public Health and Social Measures (PHSMs)

By considering the variants of concern (VOCs) and stringency index, the nonlinear infection proportionality coefficient *C*_*i*_(*t*) was defined mathematically as

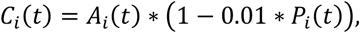

where *A*_*i*_(*t*) is the baseline of *C*_*i*_(*t*). Baseline *A*_*i*_(*t*) was estimated by Ensemble Adjustment Kalman Filter (EAKF) algorithm, and the upper bound of *A*_*i*_(*t*) was based on dominant variant during each period, details of *A*_*i*_(*t*) were provided in Supplementary Material Section 2.4.

### Parameter Estimation and Fitting Procedure

We applied the EAKF algorithm [22] to conduct model fitting, which is conducive for use with the high-dimension metapopulation model while providing satisfactory performance. In applying the EAKF, we used the daily incidence in province *i* at day *t*, 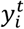, as observations, and assumed that the observation noise of incidence of COVID-19 cases followed a normal distribution with observation error variance (OEV, denoted by 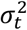)

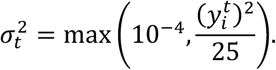

The EAKF was implemented with *n* = 500 ensemble members, assimilating observations on a daily basis. An inflation factor of 1.1 was applied to maintain ensemble spread. Details of model estimation were provided in the Supplementary Material Section 3.1.

### Heterogeneity in infection risk contribution across cluster sizes

We quantified the heterogeneity in infection risk attributable to clusters of different sizes using a Lorenz-type curve and the associated Gini coefficient. For *m*-size cluster, its contribution to the overall infection risk at time *t* in province *i* was defined as

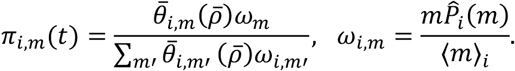

where *ω*_*i,m*_ denoted the exposure weight of *m*-size cluster in province *i*. A weighted Lorenz curve was constructed by ranking cluster sizes according to their contribution intensity per unit exposure and plotting the cumulative exposure share against the cumulative infection risk share. Deviation from the line of equality indicated unequal concentration of infection risk across cluster sizes. The corresponding Gini coefficient was computed from the area under the Lorenz curve to provide a scalar measure of heterogeneity, with larger values indicating stronger concentration of risk. The details of weighted Lorenz curve and Gini coefficient were provided in the Supplementary Material Section 4.

### Counterfactual Analysis

To quantify the impact of cluster interaction on epidemic spread, we conducted a counterfactual analysis based on the fitted model. Using identical initial conditions and parameter samples inferred from the baseline model, we simulated two scenarios:

i. Baseline scenario with the empirical cluster size distribution.
ii. Counterfactual scenario with a modified cluster size distribution 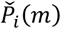 having minimized variance (Figure S6) while preserving the original mean (i.e., the average number of exposed individuals per person remains unchanged);

We compared the total number of infections across scenarios. The differences relative to the baseline quantify the effect of adjusting the cluster size distribution. Details are provided in Supplementary Material Section 5.

### Comparing the hypergraph-based SVEIRS model with the standard SVEIRS model

We hypothesized that the proposed hypergraph-based SVEIRS model could better infer the transmission dynamics of COVID-19 given a better model fit compared with the standard SVEIRS model. Thus, we also built a standard SVEIRS model with the force of infection 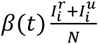, while the force of infection in hypergraph-based SVEIRS model is 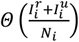. Mean Absolute Error (MAE) and Weighted Interval Score(WIS) were used as the metrics to evaluate the model fit.

### Sensitivity analysis for inter-provincial mobility and contact degree

We conducted sensitivity analyses by varying by varying the interprovincial mobility scaling factor (*δ* ± 10%) and increasing the average degree from *d* = 1.19 to *d* = 2. The details and results of several sensitivity analyses were provided in the Supplementary Material Section 6.

## Results

### Pandemic overview and temporal trends on province level in mainland China

Mainland China experienced several distinct waves during 2020-2022, each shaped by the emergence of new variants and varying levels of PHSMs. The initial outbreak began in Hubei in January 2020 and rapidly spread nationwide, marking the first major pandemic wave (Figure 1). While cases peaked in Hubei, other provinces reported limited transmission due to early containment efforts. In early February, comprehensive containment measures were implemented across provinces, which suppressed the transmission until the end of March.

**Figure 1:**
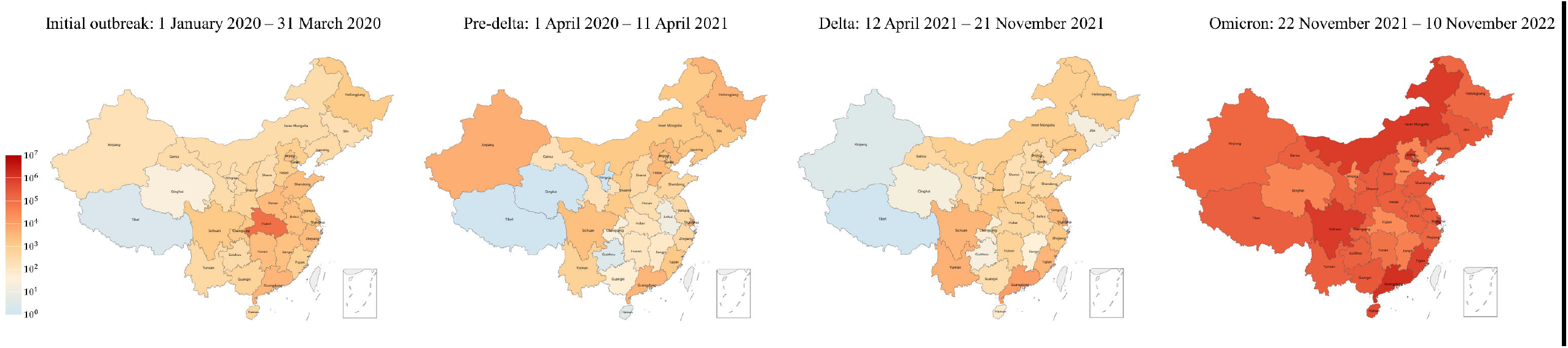
Spatial distribution of infections across provinces in mainland China at selected time points. These four periods are demarcated by the following three time points: 1 April 2020, when the first nationwide wave of the epidemic was largely brought under control; 12 April 2021, when the Delta variant emerged; and 22 November 2021, when the Omicron variant appeared.

During the per-Delta period before the emergence of the Delta variant, sporadic outbreaks occurred across the country. These were generally localized and limited in scale and duration, with higher infections observed in provinces such as Shanghai, Xinjiang, Heilongjiang, Guangdong, Hebei, and Sichuan (Figure S7). No cases reported in Tibet, Qinghai, and Ningxia (Figure S7). The spatial distribution of estimated infections in each province is shown in Figure 1.

A shift in transmission dynamics was observed following the emergence of the Delta variant in 12 April 2021. Although Delta-driven outbreaks were still rapidly contained through targeted interventions, their peak intensities generally exceeded levels observed during the pre-Delta period. These outbreaks were predominantly concentrated in southeastern coastal provinces, such as Guangdong, Shanghai, Yunnan, Fujian, and Sichuan, a major transportation hub in western China (Figure 1). Prior to the emergence of the Omicron variant on 21 November 2021, the overall number of reported cases remained relatively low.

The emergence of the Omicron variant marked a turning point in epidemic dynamics. Widespread transmission occurred across multiple provinces, often with repeated surges and significantly higher peak case numbers than those observed during the Delta period. Although some provinces temporarily brought outbreaks under control, reinfections frequently occurred due to spillovers from neighboring regions. The most severe outbreaks were concentrated in Shanghai, Guangdong, Beijing, and Sichuan (Figure 1). By August 2022, widespread transmission was observed across nearly all provinces. The model fitting results and the observed daily case data reflect the multiple waves of the outbreak as seen in mainland China, Beijing, Shanghai, Hubei, Guangdong, Sichuan (Figure 2), as well as in other provinces (Figure S7). These provinces respectively represent the capital city, a densely populated metropolitan area, the initial outbreak center, the economically central region, and an inland province.

**Figure 2:**
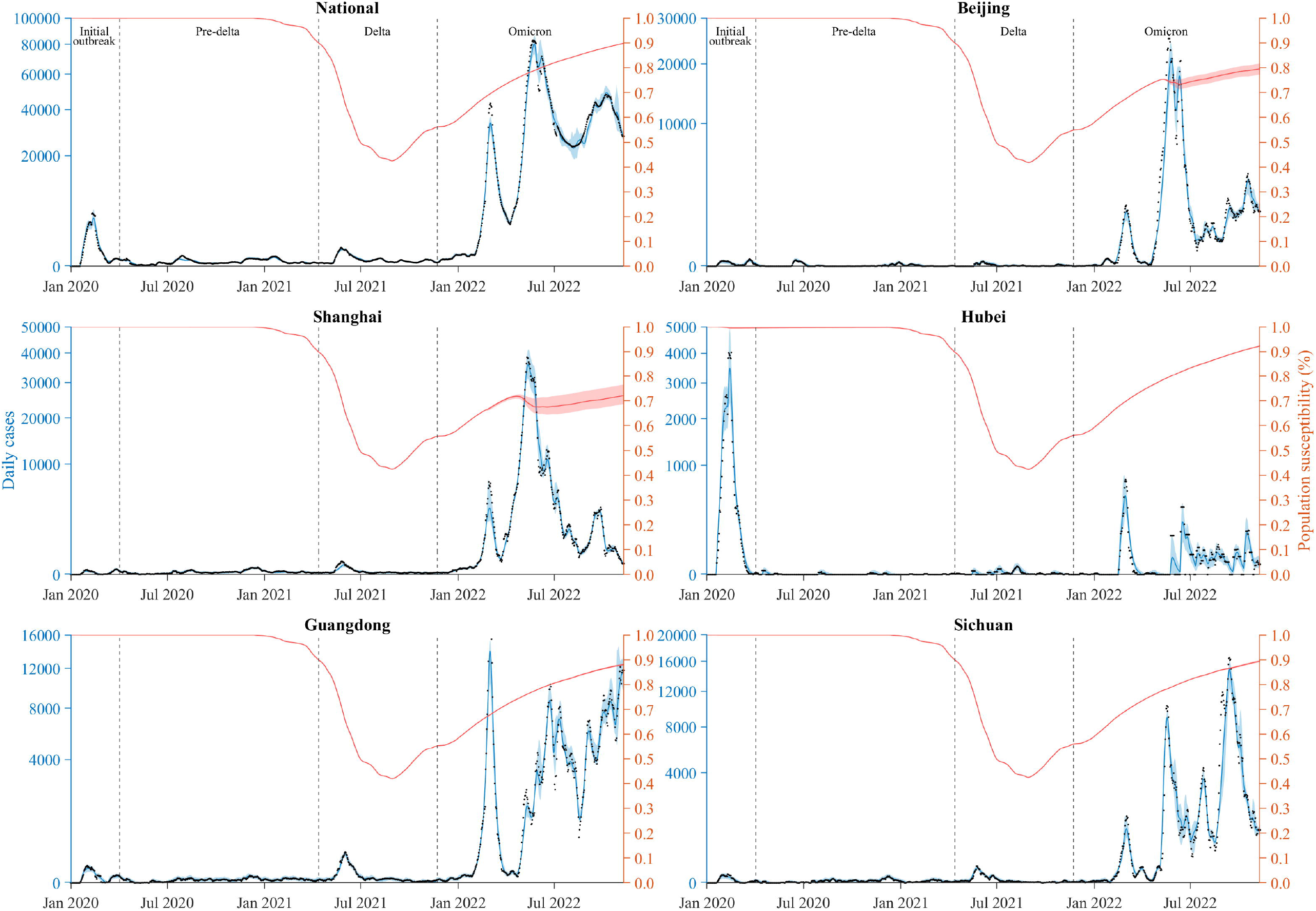
Model fitting to daily case numbers (black dots) and the susceptibility (red lines) in mainland China and five provinces (Beijing, Shanghai, Hubei, Guangdong, Sichuan) from 1 January 2020 to 10 November 2022. These provinces were selected for their epidemiological and demographic representativeness: Beijing as the national capital, Shanghai for its extremely high population density, Hubei as the initial epicenter of the COVID-19 outbreak, Guangdong as the most populous province, and Sichuan as a major transportation hub in western China. The national result was obtained by aggregating the model fit in the 31 provinces in mainland China. The blue shades represent the 95% CrI of estimated daily reported cases. The solid red lines and red shades show the median and 95%CrI of estimated susceptibility. Vertical gray dashed lines mark the beginning of the pre-Delta, Delta, and Omicron periods.

### Heterogeneity in risk contribution across cluster sizes

At both the national and provincial levels, the Lorenz curves consistently deviated from the line of equality, indicating that contributions to new infections were unevenly distributed across cluster sizes (Figure 3 and Figure S8). A relatively small fraction of total exposure accounted for a disproportionate share of new infections, demonstrating pronounced heterogeneity in transmission contributions associated with cluster size.

**Figure 3:**
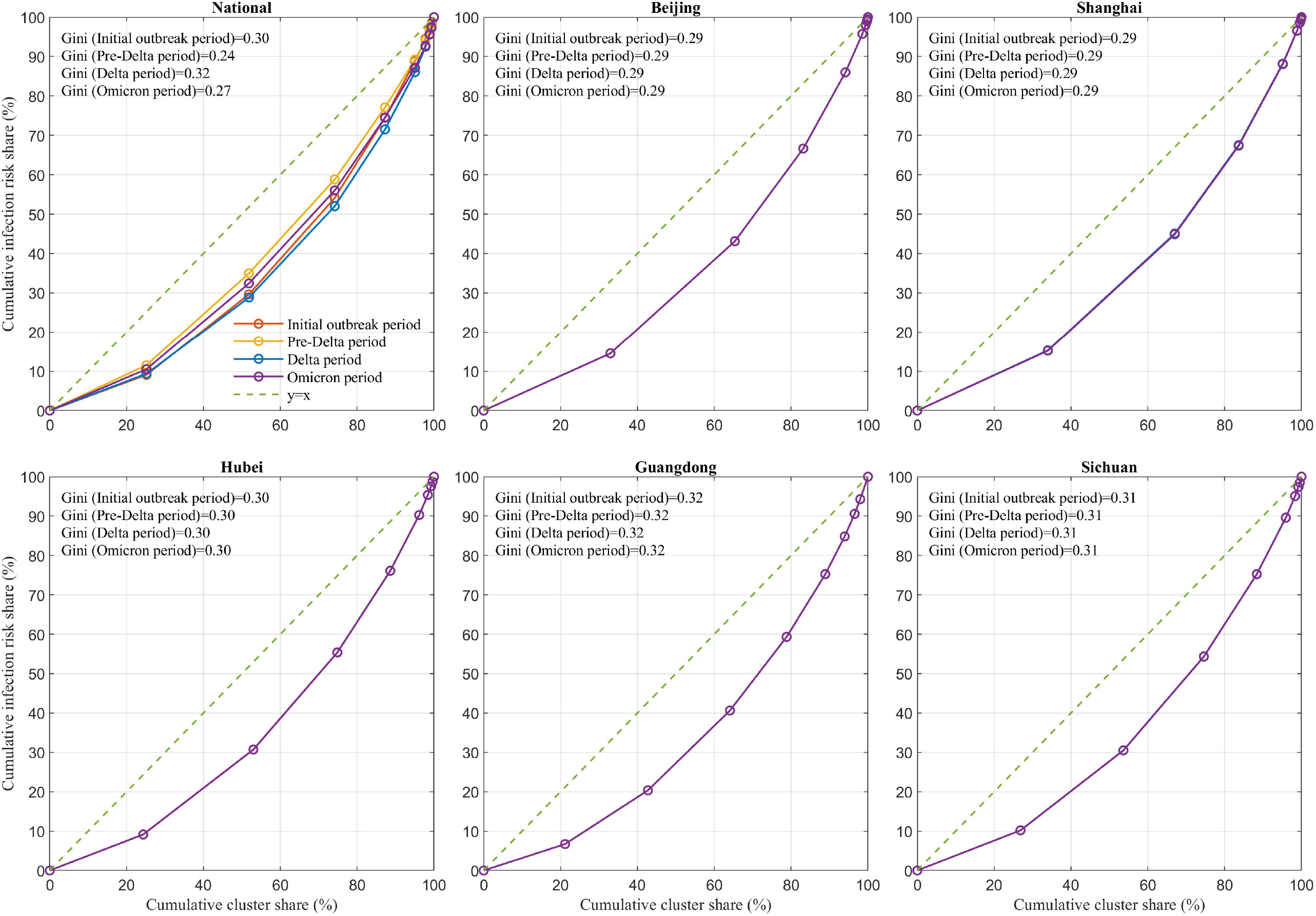
Lorenz curves depict the cumulative distribution of infection risk across cluster sizes for four study periods in mainland China and five provinces (Beijing, Shanghai, Hubei, Guangdong, Sichuan). These provinces were selected for their epidemiological and demographic representativeness: Beijing as the national capital, Shanghai for its extremely high population density, Hubei as the initial epicenter of the COVID-19 outbreak, Guangdong as the most populous province, and Sichuan as a major transportation hub in western China. The diagonal line denotes perfect equality. Gini coefficients summarize the degree of risk heterogeneity in each period.

The national Gini coefficient decreased from 0.30 during the initial outbreak period to 0.24 in the pre-Delta period, then increased to 0.32 during the Delta period before declining to 0.27 in the Omicron period (Figure 3). This non-monotonic pattern indicates substantial shifts in the concentration of infection contributions across cluster sizes over time. These changes were closely associated with the contribution patterns of large clusters. Across all periods, larger clusters consistently contributed a disproportionately high share of national infections relative to their exposure share, with the degree of disproportionality increasing with cluster size (Table 1). For example, clusters with size *m* ≥ 7 accounted for only 4.96% of total exposure but contributed between 10.77% and 14.09% of national infections across the four periods. Similarly, clusters with *m* ≥ 5, representing 25.83% of exposure, were responsible for 41.18%–47.99% of infections. The overrepresentation was even more pronounced for very large clusters: clusters with m≥10 constituted merely 0.63% of exposure, yet their contribution increased steadily from 1.65% in the initial outbreak to 2.97% during the Omicron period.

**Table 1:** Estimated contribution *π*_*m*_ (median and 95% credible interval) of clusters to national infections across epidemic periods, mainland China, 1 January 2020 – 10 November 2022.

| Cluster size group | Exposure share $\omega_m$ (%) | Initial outbreak period<br>1 Jan 2020 – 31 Mar 2020 (%) | Pre-Delta period<br>1 Apr 2020 – 11 Apr 2021 (%) | Delta period<br>12 Apr 2021 – 21 Nov 2021 (%) | Omicron period<br>22 Nov 2021 – 10 Nov 2022 (%) |
| --- | --- | --- | --- | --- | --- |
| m≥10 | 0.63 | 1.65 (1.37 - 2.19) | 1.98 (1.30 - 2.81) | 2.51 (1.89 - 3.21) | 2.97 (2.55 - 3.37) |
| m≥9 | 1.14 | 2.98 (2.57 - 3.81) | 3.37 (2.25 - 4.71) | 4.34 (3.37 - 5.44) | 4.90 (4.25 - 5.53) |
| m≥8 | 2.20 | 5.47 (4.83 - 6.76) | 5.79 (4.03 - 7.88) | 7.51 (6.06 - 9.16) | 8.05 (7.10 - 9.01) |
| m≥7 | 4.96 | 11.13 (10.06 - 13.21) | 10.77 (8.02 - 13.94) | 13.97 (11.84 - 16.33) | 14.09 (12.69 - 15.48) |
| m≥6 | 12.74 | 25.58 (24.18 - 28.22) | 22.90 (19.00 - 27.16) | 28.48 (25.56 - 31.53) | 27.35 (25.48 - 29.17) |
| m≥5 | 25.83 | 45.88 (44.05 - 48.79) | 41.18 (36.70 - 45.94) | 47.99 (44.71 - 51.35) | 46.09 (43.97 - 48.15) |
| m≥4 | 48.16 | 70.37 (68.14 - 73.49) | 65.04 (59.67 - 70.56) | 71.17 (67.26 - 75.14) | 69.51 (67.11 - 71.92) |
| m≥3 | 74.84 | 90.94 (87.36 - 94.85) | 88.46 (80.75 - 96.37) | 90.66 (85.26 - 96.09) | 90.14 (86.80 - 93.53) |
| m≥2 | 100.00 | 100.00 (95.92 - 104.36) | 100.00 (91.09 - 109.15) | 100.00 (93.85 - 106.25) | 100.00 (96.13 - 103.95) |

In contrast to the temporal variation observed at the national level, within-province heterogeneity remained remarkably stable across epidemic periods. For nearly all provinces, Gini coefficients showed minimal fluctuation across the four stages, with differences typically on the order of 10^−3^, indicating that the relative contribution structure across cluster sizes within each province was largely time-invariant (Figure S8). Despite this temporal stability, clear spatial heterogeneity was observed across provinces. Most provinces exhibited Gini coefficients within a narrow range of approximately 0.28–0.31, suggesting broadly comparable levels of transmission concentration. Nevertheless, systematic differences persisted: Inner Mongolia consistently showed lower Gini coefficients (0.27), indicating a more even distribution of infections across cluster sizes, whereas provinces such as Guangdong and Tibet exhibited higher Gini coefficients (0.32), reflecting a stronger concentration of transmission within larger clusters.

### Counterfactual effects of cluster size distribution control

At the national level, the reduction in total infections increased steadily over time, from 11.70% (95% CrI: 1.03%–21.32%) in the initial outbreak period to 14.83% (95% CrI: 12.32%–28.45%) in pre-Delta period and 27.64% (95% CrI: 10.26%–42.17%) in Delta period, reaching 30.79% (95% CrI: 20.58%– 43.72%) in Omicron period (Table 2). Most provinces broadly followed the national pattern, with relatively small reductions in the early periods and larger reductions in Omicron period. Several provinces, such as Jiangsu, Shandong, Henan, and Hubei, exhibited consistently positive effects across periods, with effects that were stable or increased over time.

**Table 2:** Provincial COVID-19 infection reduction proportion (median and 95% credible interval): minimized variance cluster size distribution 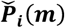 with constant per capita exposure, mainland China, 1 January 2020 – 10 November 2022.

|  | **Initial outbreak period**<br>**1 Jan 2020 – 31 Mar 2020 (%)** | **Pre-Delta period**<br>**1 Apr 2020 – 11 Apr 2021 (%)** | **Delta period**<br>**12 Apr 2021 – 21 Nov 2021 (%)** | **Omicron period**<br>**22 Nov 2021 – 10 Nov 2022 (%)** |
| --- | --- | --- | --- | --- |
| **National** | 11.70 (8.50 - 14.51) | 14.83 (9.08 - 18.88) | 27.64 (13.46 - 42.13) | 30.79 (21.02 - 34.79) |
| **Beijing** | 9.41 (6.08 - 16.67) | 11.75 (9.99 - 13.57) | 3.64 (1.39 - 6.84) | 23.35 (9.57 - 33.83) |
| **Tianjin** | 4.81 (2.67 - 9.02) | 10.60 (8.63 - 16.66) | 14.38 (9.14 - 16.57) | 19.17 (16.52 - 24.42) |
| **Hebei** | 17.89 (14.18 - 20.45) | 23.21 (11.11 - 27.79) | 5.83 (0.92 - 30.00) | 21.20 (10.75 - 26.31) |
| **Shanxi** | 6.01 (2.16 - 22.13) | 6.35 (3.84 - 13.93) | 9.44 (-3.10 - 12.66) | 22.97 (10.93 - 29.83) |
| **Inner Mongolia** | 6.68 (3.05 - 10.48) | 3.06 (1.24 - 11.11) | 2.97 (-0.55 - 5.62) | 9.09 (2.55 - 16.45) |
| **Liaoning** | 9.76 (7.07 - 16.16) | 5.31 (2.30 - 9.11) | 4.22 (1.83 - 7.68) | 6.11 (3.42 - 12.52) |
| **Jilin** | 9.05 (4.98 - 21.36) | 8.71 (-0.14 - 13.47) | 0.00 (0.00 - 100.00) | 9.43 (5.65 - 11.89) |
| **Heilongjiang** | 5.51 (-8.72 - 8.77) | 3.57 (-2.44 - 8.17) | 6.39 (1.44 - 10.51) | 8.96 (0.05 - 11.69) |
| **Shanghai** | 8.61 (4.73 - 18.41) | 12.31 (10.78 - 21.18) | 21.59 (13.51 - 29.27) | 21.40 (17.46 - 32.30) |
| **Jiangsu** | 24.73 (13.42 - 28.57) | 8.13 (5.08 - 9.66) | 25.88 (19.20 - 28.25) | 25.42 (16.72 - 35.08) |
| **Zhejiang** | 15.28 (9.55 - 25.32) | 10.89 (8.93 - 12.82) | 6.54 (3.51 - 11.07) | 14.86 (10.28 - 19.67) |
| **Anhui** | 26.68 (22.71 - 31.63) | 0.00 (0.00 - 100.00) | 2.69 (-1.29 - 10.63) | 13.49 (9.97 - 19.99) |
| **Fujian** | 24.38 (15.74 - 29.97) | 11.74 (6.10 - 14.22) | 12.01 (6.02 - 16.01) | 23.48 (7.65 - 60.09) |
| **Jiangxi** | 10.63 (0.50 - 18.20) | 6.73 (2.38 - 9.77) | 0.00 (0.00 - 100.00) | 15.42 (12.46 - 19.58) |
| **Shandong** | 16.90 (2.40 - 20.62) | 9.98 (7.61 - 13.03) | 7.66 (1.49 - 12.86) | 18.38 (14.59 - 23.08) |
| **Henan** | 26.99 (23.19 - 29.26) | 19.22 (14.76 - 28.79) | 12.51 (9.04 - 20.38) | 23.41 (13.81 - 26.93) |
| **Hubei** | 9.97 (4.41 - 12.92) | 20.84 (13.67 - 28.92) | 10.59 (5.69 - 21.43) | 11.77 (9.73 - 15.41) |
| **Hunan** | 31.02 (25.39 - 38.51) | 16.67 (12.90 - 29.41) | 9.81 (5.92 - 12.03) | 19.06 (15.15 - 22.37) |
| **Guangdong** | 30.86 (23.94 - 36.11) | 27.22 (21.85 - 31.74) | 42.58 (31.79 - 53.65) | 47.90 (20.05 - 73.10) |
| **Guangxi** | 14.47 (3.82 - 19.37) | 6.45 (-2.78 - 12.50) | 6.52 (4.10 - 18.02) | 25.17 (14.52 - 39.54) |
| **Hainan** | 24.26 (16.55 - 26.25) | 0.00 (0.00 - 0.00) | 9.26 (4.36 - 25.29) | 15.67 (11.58 - 21.07) |
| **Chongqing** | 20.31 (18.06 - 27.49) | 8.07 (0.00 - 12.90) | 7.14 (2.20 - 30.00) | 13.65 (8.85 - 21.43) |
| **Sichuan** | 22.44 (18.84 - 28.88) | 7.77 (5.37 - 11.80) | 11.80 (8.00 - 18.47) | 26.95 (21.32 - 52.28) |
| **Guizhou** | 18.55 (14.21 - 27.51) | 0.00 (0.00 - 0.00) | 6.25 (0.00 - 14.29) | 12.25 (8.04 - 15.97) |
| **Yunnan** | 16.62 (13.11 - 21.83) | 7.97 (4.96 - 9.72) | 16.22 (8.54 - 19.52) | 44.24 (22.67 - 54.06) |
| **Tibet** | 0.00 (0.00 - 0.00) | 0.00 (0.00 - 0.00) | 0.00 (0.00 - 0.00) | 21.43 (14.38 - 23.04) |
| **Shaanxi** | 17.48 (10.85 - 22.15) | 8.89 (5.34 - 11.65) | 5.52 (4.15 - 12.10) | 11.21 (3.78 - 23.82) |
| **Gansu** | 7.90 (4.89 - 9.88) | 9.87 (7.65 - 13.53) | 12.73 (6.97 - 23.34) | 31.74 (22.06 - 35.46) |
| **Qinghai** | 0.00 (-50.00 - 0.00) | 0.00 (0.00 - 0.00) | 0.00 (0.00 - 100.00) | 9.94 (4.70 - 13.25) |
| **Ningxia** | 7.56 (2.72 - 21.53) | 0.00 (0.00 - 0.00) | 10.28 (2.22 - 13.43) | 10.55 (6.03 - 16.20) |
| **Xinjiang** | 4.18 (1.53 - 14.21) | 12.38 (7.70 - 19.16) | 0.00 (0.00 - 0.00) | 17.37 (4.47 - 19.76) |

Deviations from the national trend were observed in a limited number of regions (Table 2). Beijing and Hebei showed relatively large reductions in the initial outbreak period, followed by weaker or uncertain effects in pre-Delta and Delta period, and a renewed increase in Omicron period. Shanghai exhibited a pronounced reduction in pre-Delta period, rather than a gradual increase across periods. Guangdong displayed substantial reductions already in the early periods, which continued to increase through Delta and Omicron period. In contrast, provinces such as Inner Mongolia, Liaoning, and Jilin showed persistently small or uncertain effects across most periods.

### Ascertainment rate and infections

Using the fitted model, we estimated ascertainment rates across mainland China and provinces (Figures S9). Results revealed severe under-ascertainment early in the epidemic, followed by a gradual but spatially heterogeneous improvement in detection, alongside pronounced temporal shifts in infection burden.

National ascertainment increased from 14.40% (95% CrI: 10.78%–22.69%) in January 2020 to 24.85% (95% CrI: 20.87%–29.23%) by April 2020, and rose further during the Delta period to 39.44% (95% CrI: 35.87%–43.48%) (Figure S9). By November 2022, during the Omicron period, ascertainment reached 71.93% (95% CrI: 66.48%–76.72%). Improvements were uneven across regions, with higher ascertainment in provinces such as Heilongjiang and Guangdong, and persistently low levels in Tibet and Ningxia (Figure S9).

Overall, 96.72% of all estimated infections occurred during the Omicron period, compared with 2.47% in January–March 2020 and less than 1% between April 2020 and October 2021, underscoring the strong temporal variation in both transmission intensity and detection capacity over the course of the epidemic (Figure S10).

### Temporal population immunity from infections and vaccinations

Estimated population susceptibility to SARS-CoV-2 remained extremely high at the national level during the early period of the epidemic. By 31 March 2020, national susceptibility was 99.98%, indicating that only a negligible fraction of the population had been infected, a pattern that was broadly consistent across provinces (Figure 2). Prior to the rollout of large-scale vaccination on 14 December 2020, susceptibility remained close to 100% nationwide. Following mass vaccination, susceptibility declined substantially, reaching a national minimum of 42.49% by 29 August 2021, before increasing again due to waning immunity (Figure 4). Just before the Omicron period, national susceptibility had risen to 56.03%, and by 10 November 2022, immediately prior to the implementation of the “20 Measures” policy, an estimated 89.89% of the population nationwide remained uninfected.

**Figure 4:**
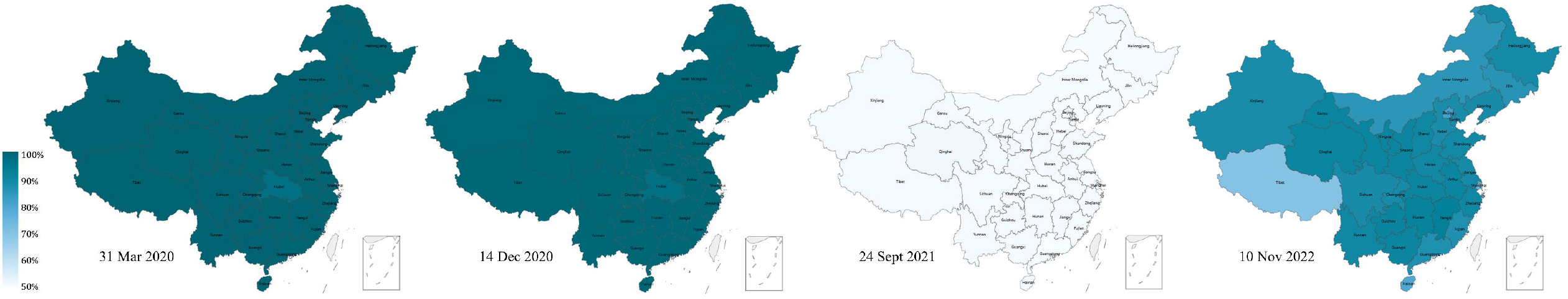
Spatial distribution of susceptibility across provinces in mainland China at selected time points. These four time points, 31 March 2020, 14 December 2020, 14 September 2021, and 10 November 2022, correspond to the end of the first epidemic wave, the period immediately prior to the implementation of nationwide COVID-19 vaccination, the time when the proportion of susceptible individuals reached its minimum, and the period immediately preceding the implementation of the “20 Measures,” respectively.

While most provinces closely followed this national trajectory, several exhibited distinct dynamics during periods of intense transmission (Figure S7). In particular, large outbreaks during the Omicron period led to temporary, localized reductions in susceptibility in a small number of provinces, most notably Beijing, Shanghai, Hainan, and Tibet, deviating from the otherwise steady national increase driven by immune waning.

### Effective reproduction number and assessment of the PHSMs

At the national level, the effective reproduction number *R*_*e*_ exhibited distinct temporal patterns across the four epidemic periods. During the initial outbreak period, national *R*_*e*_ declined from an early value of 1.69 (95% CrI: 1.42–1.98) to 0.88 (95% CrI: 0.73–1.04) by 26 January 2020 and remained below 1.0 thereafter (Figure 5). Over the same period, the national stringency index increased from 0.85 to 51.34 (Figure S11). In the pre-Delta period, national *R*_*e*_ remained below 1.0 for most of the time, with relatively small fluctuations; both its overall level and temporal variability were substantially lower than those observed during the initial outbreak period. During the Delta period, the national mean and peak values of *R*_*e*_ increased moderately compared with the pre-Delta period. Although *R*_*e*_ stayed close to 1.0 for most of the time, more pronounced intermittent peaks were observed. In the Omicron period, national *R*_*e*_ displayed the highest variability among all periods, with larger peak values and sustained intervals in which *R*_*e*_ exceeded 1.0, interspersed with episodes of rapid decline.

**Figure 5:**
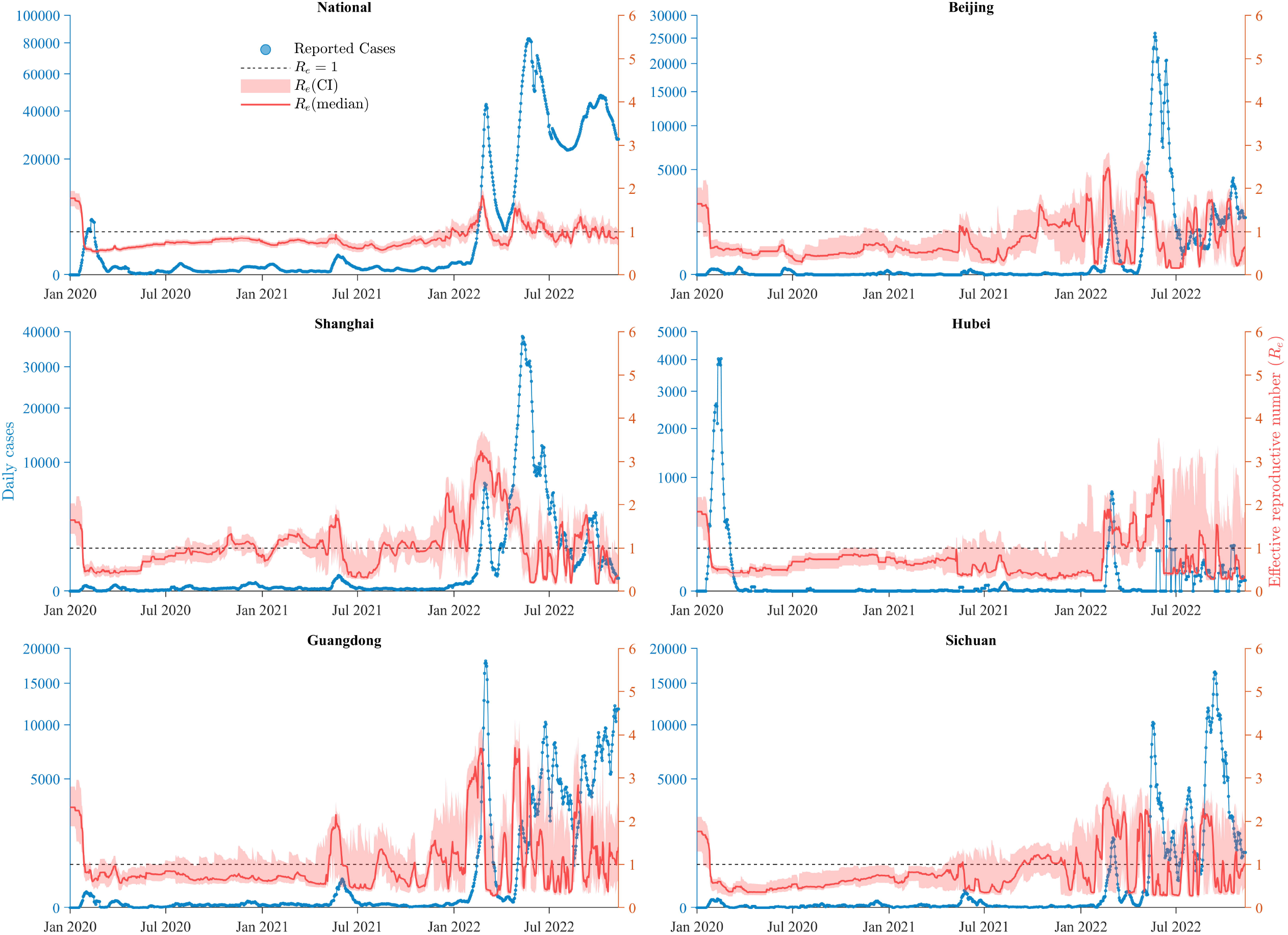
Effective reproduction number *R*_*e*_ (right y axis) and daily confirmed cases (7-d average values, left y axis) in mainland China and five provinces (Beijing, Shanghai, Hubei, Guangdong, Sichuan) from 1 January 2020 to 10 November 2022. These provinces were selected for their epidemiological and demographic representativeness: Beijing as the national capital, Shanghai for its extremely high population density, Hubei as the initial epicenter of the COVID-19 outbreak, Guangdong as the most populous province, and Sichuan as a major transportation hub in western China. The national *R*_*e*_ is calculated by the population-weight force of infection and the national cluster distribution.

At the provincial level, temporal variations in *R*_*e*_ broadly followed the national pattern across all four periods, while substantial heterogeneity persisted across provinces. During the pre-Delta period, *R*_*e*_ in most provinces fluctuated mildly around 1.0; however, Guangdong consistently exhibited higher values than the national average, reaching a peak of 1.57 on 14 December 2020 (Figure 5). In the Delta period, several provinces, including Guangdong, Shanghai, and Jiangsu, displayed sustained elevations in *R*_*e*_. Notably, Guangdong reached a *R*_*e*_ of 2.34 on 11 October 2021 when its stringency index was 45.31, after which *R*_*e*_ declined to below 1.0 following subsequent intensification of control measures. During the Omicron period, provincial heterogeneity became more pronounced. Repeated surges in *R*_*e*_ were observed in southeastern coastal regions, with Guangdong peaking at 3.86 on 27 April 2022. In addition, Jiangxi and Guangxi, despite reporting relatively fewer cases, exhibited high *R*_*e*_ values during the Omicron period, reaching 3.53 (5 August 2022) and 3.59 (30 August 2022), respectively; during these peaks, the stringency indices in both provinces remained below the national average (Figure S12).

### Performance of the proposed model over standard SVEIRS model

Table 3 presents a comprehensive comparison between the hypergraph-based model and the standard model in terms of both MAE and WIS. Overall, the hypergraph-based model consistently outperforms the standard model across most regions, demonstrating improvements in both point prediction accuracy and probabilistic forecasting quality.

**Table 3:** Performance comparison (MAE and WIS) between hypergraph-based model and standard model.

|  | **Hypergraph-based (MAE)** | **Standard (MAE)** | **Improvements% (MAE)** | **Hypergraph-based (WIS)** | **Standard (WIS)** | **Improvements% (WIS)** |
| --- | --- | --- | --- | --- | --- | --- |
| **National** | 563.2 | 1258.14 | 55.24 | 334.87 | 828.20 | 59.57 |
| **Beijing** | 160.54 | 451.71 | 64.46 | 122.91 | 402.15 | 69.44 |
| **Tianjin** | 41.48 | 45.48 | 8.79 | 22.97 | 29.05 | 20.92 |
| **Hebei** | 10.84 | 10.89 | 0.46 | 4.58 | 5.15 | 11.12 |
| **Shanxi** | 38.94 | 49.63 | 21.54 | 17.48 | 25.94 | 32.62 |
| **Inner Mongolia** | 133.93 | 136.82 | 2.11 | 76.85 | 84.30 | 8.84 |
| **Liaoning** | 30.19 | 35.13 | 14.06 | 17.22 | 20.65 | 16.62 |
| **Jilin** | 27.43 | 28.24 | 2.86 | 13.97 | 15.70 | 11.07 |
| **Heilongjiang** | 27.1 | 29.28 | 7.45 | 15.43 | 18.50 | 16.60 |
| **Shanghai** | 135.25 | 168.85 | 19.9 | 70.92 | 86.16 | 17.68 |
| **Jiangsu** | 31.51 | 33.73 | 6.56 | 15.60 | 17.67 | 11.74 |
| **Zhejiang** | 25.59 | 31.39 | 18.5 | 13.05 | 15.50 | 15.77 |
| **Anhui** | 27.81 | 38.17 | 27.15 | 11.37 | 19.88 | 42.81 |
| **Fujian** | 70.38 | 188.02 | 62.57 | 41.03 | 141.65 | 71.03 |
| **Jiangxi** | 10 | 13.24 | 24.47 | 4.75 | 6.81 | 30.16 |
| **Shandong** | 58.83 | 64.4 | 8.65 | 31.66 | 45.22 | 29.98 |
| **Henan** | 45.74 | 52.57 | 12.99 | 29.22 | 33.98 | 14.01 |
| **Hubei** | 22.38 | 23.47 | 4.63 | 10.97 | 11.62 | 5.62 |
| **Hunan** | 11.82 | 12.76 | 7.36 | 5.80 | 5.99 | 3.04 |
| **Guangdong** | 140.54 | 175.31 | 19.83 | 82.20 | 109.82 | 25.15 |
| **Guangxi** | 34.98 | 39.83 | 12.19 | 20.30 | 20.21 | -0.43 |
| **Hainan** | 48.77 | 91.8 | 46.88 | 24.85 | 40.26 | 38.27 |
| **Chongqing** | 21.9 | 23.58 | 7.1 | 11.83 | 13.17 | 10.20 |
| **Sichuan** | 123.38 | 372.09 | 66.84 | 89.90 | 324.01 | 72.25 |
| **Guizhou** | 41.49 | 36.76 | -12.89 | 23.48 | 21.86 | -7.42 |
| **Yunnan** | 28.45 | 36.56 | 22.18 | 14.06 | 18.93 | 25.74 |
| **Tibet** | 28.24 | 21.08 | -33.94 | 9.59 | 11.61 | 17.45 |
| **Shaanxi** | 29.81 | 36.17 | 17.58 | 15.22 | 18.19 | 16.30 |
| **Gansu** | 22.76 | 26.22 | 13.21 | 13.49 | 17.13 | 21.24 |
| **Qinghai** | 8.18 | 8.98 | 8.96 | 5.40 | 5.43 | 0.67 |
| **Ningxia** | 2.97 | 2.86 | -3.88 | 1.85 | 1.79 | -3.35 |
| **Xinjiang** | 22.99 | 16.65 | -38.04 | 10.98 | 8.51 | -28.96 |

At the national level, the MAE is substantially reduced from 1258.14 to 563.20, corresponding to a 55.24% decrease in prediction error. Similarly, the WIS decreases from 828.20 to 334.87, yielding a 59.57% improvement. At the provincial level, the majority of regions exhibit consistent improvements in both MAE and WIS. Significant reductions are observed in several provinces, including Beijing (MAE: 64.46%, WIS: 69.44%), Fujian (MAE: 62.57%, WIS: 71.03%), Sichuan (MAE: 66.84%, WIS: 72.25%), and Hainan (MAE: 46.88%, WIS: 38.27%). Moderate yet consistent gains are also observed in provinces such as Anhui (MAE: 27.15%, WIS: 42.81%), Jiangxi (MAE: 24.47%, WIS: 30.16%), and Yunnan (MAE: 22.18%, WIS: 25.74%). A few regions (e.g., Guizhou and Ningxia) exhibit slight degradation, while Tibet and Xinjiang show mixed results, likely due to regional heterogeneity and higher variability. Overall, the hypergraph-based model achieves robust improvements across most provinces, with reductions in both MAE and WIS demonstrating its effectiveness in capturing epidemic dynamics and associated uncertainties.

### Sensitivity analysis for *inter-provincial mobility and contact degree*

The MAE, Gini coefficient, and temporal patterns of *R*_*e*_ remained robust across all epidemic periods, although higher average degree led to larger *R*_*e*_ peaks, with corresponding model fits shown in Table S3 and Figures S13–S15.

## Discussion

Beyond structural heterogeneity, our modeling framework reconstructed the spatiotemporal dynamics of SARS-CoV-2 transmission across 31 provinces in mainland China from January 2020 to November 2022, revealing substantial variation in epidemic timing, magnitude, and detection. The majority of infections (96.72%) occurred during the Omicron period, whereas earlier waves contributed only a small fraction of the cumulative national burden (Figure 1 and Figure S10). Subsequent outbreaks remained sporadic until the emergence of the Delta variant, which expanded the spatial footprint of transmission without producing a large national surge in infections. In contrast, the Omicron variant marked a fundamental shift in epidemic dynamics, characterized by sustained increases in effective reproduction numbers and infection burden despite continued public health and social measures (Figure 5).

This study demonstrates pronounced heterogeneity in infection risk across cluster sizes. Although large clusters represent a small share of total exposure, their contributions to infections are disproportionately high and increase across epidemic phases, particularly during the Delta and Omicron periods (Table 1). Nevertheless, transmission was not driven solely by very large clusters: clusters of moderate size were the dominant contributors nationwide, with clusters of size *m* ≥ 4 consistently accounting for around two-thirds or more (65.04% - 71.17%) of infections (Table 1). This indicates that epidemic spread was sustained by a broad spectrum of clustered contacts, with large clusters acting as amplifiers rather than exclusive drivers.

Province-level Gini coefficients remained stable at approximately 0.3 because they capture the relative concentration of transmission contributions rather than differences in the underlying cluster size distributions or overall epidemic intensity (Figure S8). Across provinces, transmission was typically driven by medium-sized clusters, with larger clusters providing supplementary but non-dominant contributions. The normalization inherent in the provincial Gini calculation further dampened the influence of scale differences in case numbers, leading to similar Gini values despite substantial heterogeneity in cluster structures and epidemic trajectories.

In contrast, national Gini dynamics primarily reflected shifts in the spatial distribution of transmission across provinces rather than changes in within-province contact heterogeneity (Figure 3). Lower national Gini values during the pre-Delta (0.24) and Omicron periods (0.27) indicate more geographically widespread transmission, whereas higher values correspond to periods of spatial concentration in fewer provinces with similar cluster profiles. Counterfactual analyses further show that the epidemiological impact of cluster-size heterogeneity increased over successive phases (Table 2), with the infection reduction proportion reaching its maximum during the Delta (27.64%) and Omicron (30.79%) periods, highlighting that clustered-contact heterogeneity increasingly modulated epidemic dynamics and the responsiveness of the effective reproduction number to targeted interventions.

However, the ascertainment rate was estimated to increase continuously over time, with substantially high during the Omicron period. This trend is broadly consistent with the progressive strengthening of surveillance capacity and testing strategies in China. Testing policies were gradually expanded, extending from a narrowly defined group of symptomatic individuals to broader population-based screening during the Omicron period, including asymptomatic infections, while laboratory testing capacity and reporting systems were also improved. [16] (Figure S9). Li et al. investigated undocumented COVID-19 infections in mainland China during the early stage of the outbreak and estimated that 86% of all infections were unreported prior to the implementation of travel restrictions on 23 January 2020 [21]. Our findings are consistent with this earlier study: we estimated that 85.71% of infections were unreported in January 2020.

Vaccination substantially altered susceptibility dynamics. Following the nationwide rollout beginning in late 2020, population susceptibility declined to its lowest level in late 2021 (42.49%), which may partly explain the limited epidemic impact observed during the Delta period (Figure S7). However, this decline was followed by a rebound associated with waning immunity, particularly during the Omicron wave. By November 2022, the majority of the population remained susceptible (89.89%), reflecting both the effectiveness of prolonged containment strategies in limiting natural infection and the resulting vulnerability to large-scale outbreaks once control measures were relaxed. A seroprevalence study during April 2020 reported age- and sex-standardized seroprevalence rates of 2.66% in Wuhan, 0.033% in Shenzhen, and 0.0028% in Shijiazhuang [23]. In comparison, our model-based estimates of cumulative infection prevalence as of 30 April 2020 were 0.42% in Hubei, 0.0023% in Guangdong, and 0.0034% in Hebei.

A previous study of Wuhan in early 2020 reported a marked decline in transmission, with the effective reproduction number decreasing from about 2.35 before travel restrictions to near 1.0 afterward [24]. Our estimates for Hubei Province show a highly consistent pattern, with *R*_*e*_ declining from 1.80 to 0.97 over the same period, supporting the effectiveness of early containment measures (Figure 5). Similarly, a study of the 2021 outbreak in Guangzhou found that *R*_*e*_ dropped rapidly from very high levels (6.83) to below 1.0 within two weeks [25]. At the provincial scale, our estimates for Guangdong were lower (2.04) in magnitude but showed the same rapid decline, likely reflecting differences in spatial scale while capturing similar epidemic dynamics (Figure 5). Compared with studies focusing on transmission overdispersion inferred from detailed contact tracing and reconstructed transmission chains, the degree of transmission unevenness estimated here is lower (Table 1). This difference likely reflects the use of household-based cluster size distributions that do not capture very large clusters, as well as the absence of person-level transmission chains, making our estimates less sensitive to rare but extreme superspreading events and more representative of population-level transmission patterns [6].

This study has several limitations. The cluster size distribution is assumed to be time-invariant within each province and is approximated using household size data, which may not fully capture temporal changes in gathering patterns in non-household settings. In addition, the model represents contact heterogeneity at a distributional level rather than explicitly constructing individual or cluster interaction networks, which constrains the analysis of detailed transmission pathways and superspreading events. Finally, parameter estimation relies on data assimilation and may partially absorb unmodeled behavioural or reporting effects, highlighting the need for further investigation of parameter identifiability and complementary inference approaches in future work.

Overall, our findings indicated that the spatiotemporal heterogeneity of COVID-19 transmission in China was shaped by the interplay of viral evolution, public health interventions, immunity dynamics, population structure, and health system capacity. The prominent role of cluster size heterogeneity, and its scale-dependent manifestation, highlights the importance of incorporating structured interaction patterns into transmission models and of interpreting inequality metrics in a spatially explicit manner. These insights were critical for developing data-driven, geographically tailored strategies to manage future pandemics and the long-term transition of SARS-CoV-2 toward endemicity.

## Data Availability

All data produced in the present work are contained in the manuscript

## Notes

### Competing Interest Statement

The authors have declared no competing interest.

### Funding Statement

Health and Medical Research Fund 22210582 and 24230712 (D.W. and S.T.A.). Hong Kong Special Administrative Region, Research Grants Council 17100225 (S.T.A).

## References

1. In press. Factbox: Timeline of China’s fight against the novel coronavirus - Xinhua | English.news.cn. See http://www.xinhuanet.com/english/2020-03/19/c_138895359.htm (accessed on 30 December 2025).

2. Goldberg EE, Lin Q, Romero-Severson EO, Ke R. 2023 Swift and extensive Omicron outbreak in China after sudden exit from ‘zero-COVID’ policy. Nat. Commun. 14, 3888. (doi:10.1038/s41467-023-39638-4)

3. Adam DC, Wu P, Wong JY, Lau EHY, Tsang TK, Cauchemez S, Leung GM, Cowling BJ. 2020 Clustering and superspreading potential of SARS-CoV-2 infections in Hong Kong. Nat. Med. 26, 1714–1719. (doi:10.1038/s41591-020-1092-0)

4. Endo A, Abbott S, Kucharski A, Funk S. 2020 Estimating the overdispersion in COVID-19 transmission using outbreak sizes outside China [version 3; peer review: 2 approved]. Wellcome Open Res. 5. (doi:10.12688/wellcomeopenres.15842.3)

5. Bi Q et al. 2020 Epidemiology and transmission of COVID-19 in 391 cases and 1286 of their close contacts in Shenzhen, China: a retrospective cohort study. Lancet Infect. Dis. 20, 911–919. (doi:10.1016/S1473-3099(20)30287-5)

6. Chen D et al. 2025 Investigating setting-specific superspreading potential and generation intervals of COVID-19 in Hong Kong. Nat. Commun. 16, 5816. (doi:10.1038/s41467-025-60591-x)

7. Vyas N et al. 2023 SARS-CoV-2 transmission risk for common group activities and settings: a living scoping review. Eur. J. Public Health 34, 196–201. (doi:10.1093/eurpub/ckad195)

8. Ferretti L et al. 2024 Digital measurement of SARS-CoV-2 transmission risk from 7 million contacts. Nature 626, 145–150. (doi:10.1038/s41586-023-06952-2)

9. Thompson HA et al. 2021 Severe Acute Respiratory Syndrome Coronavirus 2 (SARS-CoV-2) Setting-specific Transmission Rates: A Systematic Review and Meta-analysis. Clin. Infect. Dis. 73, e754–e764. (doi:10.1093/cid/ciab100)

10. Iacopini I, Petri G, Barrat A, Latora V. 2019 Simplicial models of social contagion. Nat. Commun. 10, 2485. (doi:10.1038/s41467-019-10431-6)

11. Wang D, Zhao Y, Luo J, Leng H. 2021 Simplicial SIRS epidemic models with nonlinear incidence rates. Chaos Interdiscip. J. Nonlinear Sci. 31, 053112. (doi:10.1063/5.0040518)

12. St-Onge G, Sun H, Allard A, Hébert-Dufresne L, Bianconi G. 2021 Universal nonlinear infection kernel from heterogeneous exposure on higher-order networks. Phys. Rev. Lett. 127, 158301. (doi:10.1103/PhysRevLett.127.158301)

13. Dong E, Du H, Gardner L. 2020 An interactive web-based dashboard to track COVID-19 in real time. Lancet Infect. Dis. 20, 533–534. (doi:10.1016/S1473-3099(20)30120-1)

14. World Health Organization. 2023 WHO Coronavirus (COVID-19) Dashboard > Cases.

15. Our World in Data. 2023 Coronavirus Pandemic (COVID-19).

16. Hale T et al. 2021 A global panel database of pandemic policies (Oxford COVID-19 Government Response Tracker). Nat. Hum. Behav. 5, 529–538. (doi:10.1038/s41562-021-01079-8)

17. Baidu Qianxi. 2025 Baidu Migration Data Platform.

18. Wang C, Yan J. 2021 An Inversion of the Constitution of the Baidu Migration Scale Index. J. Univ. Electron. Sci. Technol. China 50, 616–626. (doi:10.12178/1001-0548.2020441)

19. Zhao Y et al. 2022 Quantifying human mixing patterns in Chinese provinces outside Hubei after the 2020 lockdown was lifted. BMC Infect. Dis. 22, 483. (doi:10.1186/s12879-022-07455-7)

20. National Bureau of Statistics of China. 2021 China Statistical Yearbook 2021. China Statistics Press. See https://www.stats.gov.cn/sj/ndsj/2021/indexeh.htm.

21. Li R, Pei S, Chen B, Song Y, Zhang T, Yang W, Shaman J. 2020 Substantial undocumented infection facilitates the rapid dissemination of novel coronavirus (SARS-CoV-2). Science 368, 489– 493. (doi:10.1126/science.abb3221)

22. Anderson JL. 2001 An ensemble adjustment kalman filter for data assimilation. Mon. Weather Rev. 129, 2884–2903. (doi:10.1175/1520-0493(2001)129<2884:AEAKFF>2.0.CO;2)

23. Chang L et al. 2021 The prevalence of antibodies to SARS-CoV-2 among blood donors in China. Nat. Commun. 12, 1383. (doi:10.1038/s41467-021-21503-x)

24. Kucharski AJ et al. 2020 Early dynamics of transmission and control of COVID-19: a mathematical modelling study. Lancet Infect. Dis. 20, 553–558. (doi:10.1016/S1473-3099(20)30144-4)

25. Li L et al. 2022 Transmission and containment of the SARS-CoV-2 Delta variant of concern in Guangzhou, China: A population-based study. PLoS Negl. Trop. Dis. 16, e0010048. (doi:10.1371/journal.pntd.0010048)

