## supplemental materials for "Transmission dynamics of the COVID-19 pandemic across the emerging variants in mainland China: a hypergraph-based spatiotemporal modeling study"

### Supplementary Material– Transmission dynamics of the COVID-19 pandemic across the emergence of dominant variants in mainland China: a hypergraph-based spatiotemporal modeling study

Yi Wang[1,2,3†], Dong Wang[1,2†], Yiu-Chung Lau[1,2†], Zhanwei Du[1,2], Benjamin J. Cowling[1,2], Yi Zhao[3,*], Sheikh Taslim Ali[1,2,*]

**Affiliations:**

[1] WHO Collaborating Centre for Infectious Disease Epidemiology and Control, School of Public Health, Li Ka Shing Faculty of Medicine, The University of Hong Kong, Hong Kong Special Administrative Region, China

[2] Laboratory of Data Discovery for Health, Hong Kong Science and Technology Park, Hong Kong Special Administrative Region, China

[3] School of Science, Harbin Institute of Technology, Shenzhen, China

† Contributed equally

*Corresponding Author:

Yi Zhao,

### 1. Data processing

#### *1.1. Surveillance data*

The JHU CSSE dataset [1] began on 22 January 2020, and the WHO dataset [2] on 4 January 2020, because the WHO dataset could change includes retrospective updates to reflect reporting corrections. To standardize the temporal coverage of the dataset and facilitate downstream analyses such as delay-adjusted reporting and early outbreak dynamics, we defined 1 January 2020 as the starting point of the study period. We padded the missing values prior to their respective start dates with zeros for all provinces. Before 31 March 2020, the officially reported COVID-19 cases in mainland China were province-specific and did not distinguish between confirmed cases (i.e., symptomatic individuals who tested positive by PCR) and asymptomatic infections (i.e., individuals without symptoms but PCR-positive). After 31 March 2020, China separately reported the province-level confirmed cases and national-level asymptomatic infections. For consistency in definitions, the JHU CSSE dataset includes only confirmed cases and excludes asymptomatic infections. In contrast, the WHO dataset includes the total number of infections in mainland China (national-level), including both confirmed and asymptomatic cases.

#### *1.2. Vaccination data*

We obtained four indicators from the vaccination dataset [3]: (1) the number of individuals who received at least one dose, (2) the number of individuals who completed the initial vaccination protocol, (3) the number of individuals who received a booster dose, and (4) the total number of vaccine doses administered. The fourth indicator represented the total number of doses (all doses, including boosters) are counted individually. All four indicators exhibited missing values to varying degrees, among which the total dose count is the most complete. Therefore, we used this indicator as the basis for imputing missing values in the other three.

The total dose data began on 15 December 2020. First, we applied linear interpolation to fill the missing values within this series. We observed that the first available values of the other three indicators were disproportionately high, likely due to the delayed start of data collection relative to the actual beginning of vaccination campaigns. To address this, second, we redefined the start dates for these three series based on documented vaccination rollout timelines (see Table S1) and linearly interpolated the missing values from the revised starting points onward.

Third, based on the imputed cumulative counts of the first dose, second dose, and booster dose, we derived the corresponding daily increments. Finally, we used the relative daily proportions of these three dose types to redistribute the total daily administered doses, thus obtaining a consistent estimate of daily new recipients for the first, second, and booster doses.

##### *1.3. Mobility data*

Baidu Qianxi provided both the migration scale index and the migration proportion; however, these data exhibited varying degrees of missingness during the study period from 1 January 2020 to 10 November 2022. Specifically, Baidu Qianxi only released data for select periods, such as the Spring Festival travel season (Chunyun), public holidays, and other peak travel times. Since the 2023 mobility data are complete, we addressed the missing values from 2020 to 2022 by aligning weekends and holidays and using the corresponding 2023 data to impute the gaps.

##### *1.4. Degree data*

To estimate the average number of clusters an individual participates in per day, we utilized contact survey data in Chinese provinces outside Hubei collected between 3 March and 23 March 2020 [4]. The survey recorded all social contacts experienced by each participant within a 24-hour period, where contacts are defined as either a face-to-face conversation involving more than three spoken words or direct physical interactions, along with the location where the contact occurred. For each participant, the number of distinct types of locations visited on the surveyed day was calculated and considered as the number of clusters the individual engaged in. The daily average number of clusters was then obtained by averaging across all participants.

#### 2. Hypergraph-based model

##### *2.1. Compartmental model for COVID-19 transmission in mainland China at province level*

We formulated the hypergraph-based SVEIRS model as the following differential equations,

$$
\begin{aligned}
\frac{dS_i(t)}{dt} &= -\Theta_i\left(\frac{I_i^r(t)+I_i^u(t)}{N_i(t)}\right)S_i(t)-\eta_i(t)+\lambda V_i(t)-b_i(t)+\gamma R_i(t)+\sum_j\frac{M_{ij}(t)S_j(t)}{N_j(t)-I_j^r(t)}-\sum_j\frac{M_{ji}(t)S_i(t)}{N_i(t)-I_i^r(t)}\\
\frac{dV_i(t)}{dt} &= -\left(1-\square\square(t)\right)\Theta_i\left(\frac{I_i^r(t)+I_i^u(t)}{N_i(t)}\right)V_i(t)+\eta_i(t)-\lambda V_i(t)+b_i(t)+\sum_j\frac{M_{ij}(t)V_j(t)}{N_j(t)-I_j^r(t)}-\sum_j\frac{M_{ji}(t)V_i(t)}{N_i(t)-I_i^r(t)}\\
\frac{dE_i(t)}{dt} &= \Theta_i\left(\frac{I_i^r(t)+I_i^u(t)}{N_i(t)}\right)\left(S_i(t)+\left(1-\varepsilon(t)\right)V_i(t)\right)-\frac{E_i(t)}{Z}+\sum_j\frac{M_{ij}(t)E_j(t)}{N_j(t)-I_j^r(t)}-\sum_j\frac{M_{ji}(t)E_i(t)}{N_i(t)-I_i^r(t)}\\
\frac{dI_i^r(t)}{dt} &= \alpha_i(t)\frac{E_i(t)}{Z}-\frac{I_i^r(t)}{D}\\
\frac{dI_i^u(t)}{dt} &= \left(1-\alpha_i(t)\right)\frac{E_i(t)}{Z}-\frac{I_i^u(t)}{D}+\sum_j\frac{M_{ij}(t)I_j^u(t)}{N_j(t)-I_j^r(t)}-\sum_j\frac{M_{ji}(t)I_i^u(t)}{N_i(t)-I_i^r(t)}\\
\frac{dR_i(t)}{dt} &= \frac{(I_i^r(t)+I_i^u(t))}{D}-\gamma R_i(t)+\sum_j\frac{M_{ij}(t)R_j(t)}{N_j(t)-I_j^r(t)}-\sum_j\frac{M_{ji}(t)R_i(t)}{N_i(t)-I_i^r(t)}\\
N_i(t) &= S_i(t)+V_i(t)+E_i(t)+I_i^r(t)+I_i^u(t)+R_i(t)
\end{aligned}
\tag{1}
$$

Here, $S_i(t)$, $V_i(t)$, $E_i(t)$, $I_i^r(t)$, $I_i^u(t)$, $R_i(t)$, and $N_i(t)$ are the susceptible, vaccinated, exposed, reported infectious, unreported infectious, recovered, and total population in province $i$ at day $t$. $\Theta_i(\cdot)$ is the hypergraph-based force of infection of each susceptible individual in province $i$, $\eta_i(t)$ represents the number of individuals transitioning from the susceptible to the vaccinated as a result of vaccination at day $t$ in province $i$, $\lambda$ represents vaccine-induced immunity waning rate, $b_i(t)$ denotes the number of individuals who receive booster doses and maintain immunity, $\varepsilon(t)$ is the vaccination effectiveness at day $t$. $\alpha_i(t)$ is the ascertainment rate in province $i$ on the day $t$, $Z$ represents the average latency period and $D$ represents the average duration of being infectious. $M_{ij}(t)=\delta M_0(t)m_{ij}(t)$ is the approximated human mobility flow from province $j$ to province $i$ at day $t$, where $\delta = 3.09 \times 10^5$ is a multiplicative scalar [5], $M_0(t)$ is the migration scale index, and $m_{ij}(t)$ is the proportion of individuals moving from location $j$ to location $i$ among the total outflow from location $j$. The heatmap illustrated the inter-provincial mobility flows $M_{ij}(t)$ across the 31 provinces in mainland China on 13 February 2020, which corresponds to the peak of the first epidemic wave (Figure S2). $\gamma$ is infection-induced immunity waning rate. We assumed the birth rate and death rate were negligible to the transmission dynamics and thus were not considered in the model [6]. The structure of SVEIRS model illustrated in Figure S3.

##### *2.2. Hypergraph-based force of infection*

Based on a hypergraph contact network, we proposed the hypergraph-based force of infection, which was nonlinearly related to infected contacts by integrating heterogeneous exposure with the concept of minimal infective dose. We ignored the province-specific notation in this section for simplicity.

Considering the cluster size, duration of participation, the minimum infectious dose of the virus, and other factors, St-Onge et al. assumed that the virus dose follows an exponential distribution and pointed out that the force of infection of a susceptible individual in an $m$-size household cluster containing $k$ infected individuals is as follows [7],

$$\theta_{i,m}(\rho) = C_i(t) \cdot (\rho \cdot f(m))^{v_i}. \quad (2)$$

Herein $C_i(t)$ is the nonlinear infection proportionality coefficient at day $t$ in province $i$, $\rho = \frac{k}{m-1}$ is the density of infected individuals in this cluster, and $f(m) = m - 1$ is the typical number of contacts in environments. $v_i > 0$ is the nonlinear kernel in province $i$ which is related to the heterogeneous exposure periods distribution and the virus dose decrease velocity, leading to a nonlinear growth for the force of infection. Illustrative examples of the force of infection to a susceptible in the $m$-size cluster are shown in Figure S4.

Assuming the number of infectious individuals in a $m$-size household cluster follows a binomial distribution with parameter $\bar{\rho}_i = \frac{I_i^r + I_i^u}{N_i}$ denoting the density of infectious individuals in the entire population, the expected force of infection to a susceptible individual in a $m$-size cluster $(\bar{\theta}_{i,m}(\bar{\rho}_i))$ was calculated as,

$$\bar{\theta}_{i,m}(\bar{\rho}_i) = \sum_{k=0}^{m-1} \binom{m-1}{k} \bar{\rho}_i{}^k (1-\bar{\rho}_i)^{m-1-k} \theta_{i,m}\left(\frac{k}{m-1}\right).$$

Given the cluster size distribution data $\hat{P}_i(m)$ of province $i$, the expectation of the individual force of infection $\bar{\theta}_i(\bar{\rho}_i)$ in a certain cluster is the average of $\bar{\theta}_{i,m}(\bar{\rho}_i)$ over the cluster size distribution,

$$\bar{\theta}_i(\bar{\rho}_i) = \sum_m \bar{\theta}_{i,m}(\bar{\rho}_i) \frac{m\hat{P}_i(m)}{\langle m \rangle_i}.$$

$\langle m \rangle_i$ is denoted as the mean cluster size of province $i$. Then, we calculated the force of infection in the contact network. Assuming that the probability of infection through each cluster is independent, we derived the hypergraph-based force of infection as,

$$\Theta_i\left(\frac{I_i^r + I_i^u}{N_i}\right) = 1 - \left(1 - \bar{\theta}_i\left(\frac{I_i^r + I_i^u}{N_i}\right)\right)^d,$$

where $d$ is the number of contact clusters. Based on the cluster data, we assume that each individual participates in $d = 1.19$ clusters per day [4]. The cluster size distribution, $\hat{P}_i(m)$, was assumed to follow the household scale distribution at the provincial level in mainland China [8], shown in Figure S5. This implies that the cluster size distributions for both the workplace and the school are also assumed to follow the same household scale distribution.

##### *2.3. Vaccination effectiveness*

We estimated the vaccination effectiveness $\varepsilon(t)$ by considering the variants of concern (VOCs). Data on SARS-CoV-2 VOCs in mainland China were obtained from CoVariants [9]. This dataset reported the proportion of viral genome sequences, rather than confirmed cases, that belong to defined variant groups over time. Sequence data are aggregated in two-week intervals, providing a temporal distribution of variant prevalence based on submitted sequencing results. Given that the COVID-19 outbreak in China spanned three major VOCs, the Ancestral, Delta, and Omicron, we found that the ancestral remained dominant until 12 April 2021, when the Delta variant emerged and gradually began to replace it. Delta became the predominant circulating variant by 19 July 2021. Subsequently, the Omicron variant was first detected on 22 November 2021, and within 14 weeks—by 28 February 2022—it had supplanted Delta as the dominant variant, maintaining this status through the end of 2022. The effectiveness of inactivated COVID-19 vaccines against Ancestral, Delta, and Omicron variants were 0.79, 0.51, and 0.27, respectively [10–12]. Therefore, we set the vaccination effectiveness as a piecewise function across five epidemic phases, based on the dominant circulating variant during each phase (Figure S1).

##### *2.4. Effect of Public Health and Social Measures (PHSMs)*

We estimated the nonlinear infection proportionality coefficient $C_i(t)$ by considering the VOCs and stringency index. Specifically, $C_i(t)$ was defined mathematically as

$$C_i(t) = A_i(t) * \left(1 - 0.01 * P_i(t)\right),$$

where $A_i(t)$ is the baseline of $C_i(t)$. The upper bound of the baseline $A_i(t)$ for province $i$ was defined as a piecewise function divided into five phases according to the dominant circulating variants. Specifically, for province $i$, the upper bound during the Ancestral period was set to $0.13 * \frac{PD_i}{PD_{median}}$, based on the reported upper bound of the basic reproduction number $R_0 = 3.44$ for the Ancestral [13]. During the Delta period, the upper bound was set to $0.30 * \frac{PD_i}{PD_{median}}$, corresponding to the upper bound $R_0 = 8.00$ for the Delta variant [14]. During the Omicron period, the upper bound was set to $0.48 * \frac{PD_i}{PD_{median}}$, based on the upper bound $R_0 = 13.00$ for the Omicron variant [15]. Here, $PD_i$ denotes the population density of province $i$, and $PD_{median}$ represents the median population density across the 31 provinces. For the transition intervals between variants (Phase 2 and 4), the upper bound of baseline $A_i(t)$ varied linearly between the corresponding constants, ensuring a smooth and continuous adjustment of upper bound over time.

##### *2.5. Hypergraph-based reproduction number*

Details of hypergraph-based basic reproductive numbers can be found in St-Onge et al. [7]. In brief, the average number of new infections caused by an infected node in a single time step at the onset of the epidemic is equal to

$$\left(\frac{\left\langle m(m-1)\theta_{i,m}\left(\frac{1}{m-1}\right)\right\rangle}{\langle m\rangle_i}\right)\left(\frac{\langle d^2\rangle}{\langle d\rangle}\right).$$

Then we obtained the effective reproduction number at day $t$ in province $i$ as,

$$R_{i,t} = D\left(\frac{\left\langle m(m-1)\theta_{i,m}\left(\frac{1}{m-1}\right)\right\rangle}{\langle m\rangle_i}\right)\left(\frac{\langle d^2\rangle}{\langle d\rangle}\right)\frac{S_i(t)+\left(1-\varepsilon(t)\right)*V_i(t)}{N},$$

##### *2.6. Reporting delay*

To account for delays in infection report, we mapped the simulated new reported infections cases $\alpha_i(t)\frac{E_i}{Z}$ to daily surveillance data with an observational delay. A prior study suggested that the reporting delay $T_d$ from 24 January 2020 to 8 February 2020, followed Gamma distribution with a mean of 6 days and a standard deviation of 19.46 days (with shape parameter of 1.85 and scale parameter of 3.24) [13]. In this delay model, the reporting delay includes the time interval between a person transitioning from latent to reported infectious (i.e., $E \rightarrow I^r$) and the reporting of that individual infection.

Given the consideration of reporting delays, at time $t$, we incorporated a $T_d$ days forward projection to estimate the daily reported cases at time $t+T_d$ using the delay observation model. The EAKF algorithm was then used to calibrate the model based on the observed daily reported cases. Essentially, we applied a delayed gamma distribution model to optimize the current model variables and parameters.

### 3. Model likelihood and estimations

#### *3.1. Ensemble Adjustment Kalman Filter (EAKF) algorithm*

We applied the EAKF [14] algorithm to estimate parameters and states in the hypergraph-based SVEIRS model. This algorithm was applied to high-dimensional models and has been successfully used to infer the epidemiological parameters for a range of infectious diseases [13,15]. EAKF is a recursive Bayesian estimation technique, assumed following normal distribution, that updates parameter estimates in real time by minimizing the mean squared error between model predictions and observed data. This algorithm updated model states and parameters by processing observations sequentially and computing each state variable independently, while accounting for process and measurement noise, making it highly efficient for application in large models. This ensemble method can assimilate observations with a nonlinear relation to model state variables.

The EAKF algorithm was implemented in MATLAB R2021a [15], with adjustments made according to the specific requirements of our epidemiological model. The EAKF algorithm updates states and parameters estimates based on daily incidence data while accounting for both model and observation uncertainties. The observation uncertainty, represented by the observation error variance (OEV, denoted by $\sigma_t^2$). A range of OEV settings were tested, and we set the standard deviation of an observation as 20% of its value,

$$\sigma_t^2 = \max\left(10^{-4}, \frac{(y_i^t)^2}{25}\right),$$

where $y_i^t$ is the daily incidence in province $i$ at day $t$ [13,15]. In this study, the 95% credible interval was computed using the 2.5th and 97.5th percentiles derived from $n$ ensemble trials. The ascertainment rate $\alpha_i(t)$ were updated per two weeks. The baseline infection coefficient $A_i(t)$ were updated daily. To reflect the continuous improvement in detection capacity, $\alpha_i(t)$ was constrained to be monotonically increasing over time. The model parameters and the corresponding priors used were summarized in Table S2.

To justify modeling the ascertainment rate as a monotonically increasing function over time, we draw support from multiple lines of evidence. First, early epidemiological investigations indicate that COVID-19 ascertainment was severely limited during the initial phase of the pandemic, with only a small fraction of infections being detected [16]. Second, testing policies in China underwent substantial expansion over the course of the pandemic: testing was initially restricted to symptomatic individuals meeting strict eligibility criteria, then gradually extended to all symptomatic persons, and eventually opened to broad population-based screening, including asymptomatic individuals [17]. Third, During the 2022 Omicron variant outbreak, actively encouraging nucleic acid testing for residents in affected areas and the immediate isolation of identified cases and their close contacts [18]. Together, these observations support the biological and policy-driven plausibility of imposing a monotonically increasing constraint on the ascertainment rate throughout the study period.

##### *3.2. Model initialization*

To initialize the model-inference system, we seeded exposed individuals ($E$) and unreported infected ($I^u$) in provinces with at least 1 confirmed cases. We randomly drew the initial $E$ and $I^u$ from uniform distributions $[0,12U]$ and $[0,10U]$ five days before the first date with more than five confirmed cases $T_0$, where $U$ is the total number of confirmed cases between day $T_0$, and $T_0 + 4$. This setting provided a broad seeding range for provinces in mainland China.

#### 4. Weighted Lorenz curve and Gini coefficient

This section provides technical details on the construction of the weighted Lorenz curve and the computation of the corresponding Gini coefficient used to quantify heterogeneity in infection risk across different cluster sizes. $\omega_{i,m} = \frac{m\hat{P}_i(m)}{\langle m \rangle_i}$ denotes the $m$-size cluster exposure weight in province $i$, representing the probability that a randomly selected individual belongs to an $m$-size cluster, where $\hat{P}_i(m)$ refers to the cluster size distribution of province $i$, and $\langle m \rangle$ denotes the average cluster size. For a $m$-size cluster, its contribution to the overall infection risk at time $t$ was defined as $\pi_{i,m}(t) = \frac{\bar{\theta}_{i,m}(\bar{\rho}_i)\omega_{i,m}}{\sum_{m'} \bar{\theta}_{i,m'}(\bar{\rho}_i)\omega_{i,m'}}$, satisfying $\sum_m \pi_{i,m}(t) = 1$.

Because cluster sizes contribute to infection risk with unequal exposure weights $\omega_m$, we adopted a weighted Lorenz formulation. Specifically, cluster sizes were ranked in ascending order according to their contribution intensity per unit exposure, defined as $r_{i,m}(t) = \pi_{i,m}(t)/\omega_{i,m}$. After reordering, the weighted Lorenz curve was constructed by plotting the cumulative exposure share $X_{i,k} = \sum_{p=1}^{k} \omega_{i,(p)}$ against the cumulative contribution share $Y_{i,k} = \sum_{p=1}^{k} \pi_{i,(p)}$, with the curve anchored at $(0,0)$ and $(1,1)$. The $\omega_{i,(p)}$ and $\pi_{i,(p)}$ are the reordered $\omega_{i,m}$ and $\pi_{i,m}$. This construction ensures that the Lorenz curve lies at or below the line of equality when contributions are nonnegative and properly ordered.

The Gini coefficient was computed from the weighted Lorenz curve as $G_i = 1 - 2\int_0^1 Y_i(X_i)dX_i$, which, in the discrete case, was evaluated using the trapezoidal rule. By definition, $G_i \in [0,1]$, where $G_i = 0$ corresponds to perfectly homogeneous contributions across cluster sizes, and larger values indicate increasing concentration of infection risk. All computations were performed using normalized $\omega_{i,m}$ and $\pi_{i,m}(t)$ to ensure numerical stability.

The $m$-size cluster exposure weight at the national level was defined as $\omega_m = \frac{m\hat{P}_i(m)}{\langle m \rangle}$, where $\hat{P}(m)$ refers to the cluster size distribution of mainland China. To obtain national-level contribution by $m$-size cluster, the cluster-size–specific contributions were first computed at the provincial level based on the decomposition of the SVEIRS infection term. Specifically, for province $i$, the absolute contribution of $m$-size clusters to new infections at time $t$ was defined as

$$H_{i,m}(t) = \bar{\theta}_{i,m}(\bar{\rho}_i)\omega_{i,m}(S_i(t) + (1 - \varepsilon(t))V_i(t)).$$

National-level contributions were then obtained by aggregating these absolute contributions across provinces, $H_m(t) = \sum_i H_{i,m}(t)$, and normalized to yield the national contribution share $\pi_m(t) = \frac{H_m(t)}{\sum_m H_m(t)}$. Based on the resulting $\pi_m(t)$ and the corresponding exposure weights $\omega_m$, the weighted Lorenz curve was constructed by plotting the cumulative exposure share against the cumulative contribution share. The associated Gini coefficient was subsequently computed as twice the area between the Lorenz curve and the line of equality.

#### 5. Counterfactual analysis

To systematically evaluate the role of cluster-size heterogeneity in transmission dynamics, we constructed a counterfactual household-scale distribution $\check{P}_i(m)$ that modify dispersion while strictly preserving the empirical mean for each province. The empirical distribution $\hat{P}_i(m)$ is defined on $m = 2,3,\ldots,10$ with empirical mean $\langle m \rangle_i$.

For province $i$, to obtain the variance-minimizing cluster size distribution $\check{P}_i(m)$, we solved the following constrained optimization problem:

$$\min_{\check{P}_i} \mathrm{Var}_{\check{P}_i}(m) \quad s.t. \sum_m \check{P}_i(m) = 1, \qquad \sum_m m\check{P}_i(m) = \langle m \rangle_i.$$

Because

$$\mathrm{Var}(m) = \mathbb{E}(m^2) - {\langle m \rangle_i}^2$$

and the mean $\langle m \rangle_i$ is fixed, minimizing variance is equivalent to minimizing $\mathbb{E}(m^2)$ under linear constraints. This is a linear objective over a convex feasible set. Therefore, the optimal solution is attained at an extreme point, which under two linear constraints (normalization and fixed mean), has at most two nonzero probability masses. For each province $i$, define

$$m_L = \lfloor \langle m \rangle_i \rfloor, \qquad m_U = \lceil \langle m \rangle_i \rceil.$$

The variance-minimizing counterfactual distribution is supported on $m_L$ and $m_U$ with weights $\omega_L$ and $\omega_U$ satisfying

$$\omega_L + \omega_U = 1, \qquad m_L\omega_L + m_U\omega_U = \langle m \rangle_i.$$

Thus,

$$\check{P}_i(m) = \begin{cases} \omega_L & m = m_L \\ \omega_U & m = m_U \\ 0 & otherwise \end{cases},$$

where

$$\omega_U = \frac{\langle m \rangle_i - m_L}{m_U - m_L}, \qquad \omega_L = 1 - \omega_U.$$

This construction guarantees: (i) exact preservation of the empirical mean $\langle m \rangle_i$, and (ii) minimal variance among all discrete distributions with the same support and mean.

The modified distribution is subsequently used in counterfactual simulations to quantify how cluster size distribution heterogeneity influences epidemic spread. The adjusted cluster size distribution of each province is shown in Figure S6.

#### 6. Sensitivity analysis

Several sensitivity analyses were performed to examine the robustness of the inference results.

1) We chose the multiplicative factor adjusting interprovincial mobility as $\delta = 3.09 \times 10^5$. We repeated the inference using alternative mobility scaling factors, $\delta_1 = 1.1\delta$ and $\delta_2 = 0.9\delta$. The mean absolute error (MAE), Gini coefficient, and peak values of the effective reproduction number $R_\mathrm{e}$ were highly robust to moderate perturbations in the mobility scaling factor across all four epidemic periods (Table S3). The model fitting compared with the baseline scenario is shown in Figures S13 and S14.
2) In the main analysis, we assumed an average degree of $d = 1.19$, corresponding to each individual participating in 1.19 clusters per time unit. We repeated the inference with $d = 2$. The MAE and Gini coefficient were robust to moderate changes in the assumed average degree. Increasing the average degree resulted in systematically higher $R_\mathrm{e}$ peaks, reflecting the direct role of contact aggregation frequency in shaping transmission intensity. Despite these shifts in magnitude, the relative temporal pattern of $R_\mathrm{e}$ across epidemic periods remained unchanged (Table S3). The model fitting compared with the baseline scenario is shown in Figure S15.

#### 7. References

1. Dong E, Du H, Gardner L. 2020 An interactive web-based dashboard to track COVID-19 in real time. *Lancet Infect. Dis.* **20**, 533–534. (doi:10.1016/S1473-3099(20)30120-1)

2. World Health Organization. 2023 WHO Coronavirus (COVID-19) Dashboard > Cases.

3. World Health Organization. 2023 WHO Coronavirus (COVID-19) Dashboard > Vaccines.

4. Zhao Y *et al.* 2022 Quantifying human mixing patterns in Chinese provinces outside Hubei after the 2020 lockdown was lifted. *BMC Infect. Dis.* **22**, 483. (doi:10.1186/s12879-022-07455-7)

5. Wang C, Yan J. 2021 An Inversion of the Constitution of the Baidu Migration Scale Index. *J. Univ. Electron. Sci. Technol. China* **50**, 616–626. (doi:10.12178/1001-0548.2020441)

6. Xiao H, Wang Z, Liu F, Unger JM. In press. Excess All-Cause Mortality in China After Ending the Zero COVID Policy.

7. St-Onge G, Sun H, Allard A, Hébert-Dufresne L, Bianconi G. 2021 Universal nonlinear infection kernel from heterogeneous exposure on higher-order networks. *Phys. Rev. Lett.* **127**, 158301. (doi:10.1103/PhysRevLett.127.158301)

8. National Bureau of Statistics of China. 2021 *China Statistical Yearbook 2021*. China Statistics Press. See https://www.stats.gov.cn/sj/ndsj/2021/indexeh.htm.

9. Hodcroft EB. 2021 CoVariants: SARS-CoV-2 Mutations and Variants of Interest.

10. Ismail AlHosani F *et al.* 2022 Impact of the Sinopharm's BBIBP-CorV vaccine in preventing hospital admissions and death in infected vaccinees: Results from a retrospective study in the emirate of Abu Dhabi, United Arab Emirates (UAE). *Vaccine* **40**, 2003–2010. (doi:10.1016/j.vaccine.2022.02.039)

11. Wu D *et al.* 2022 Effectiveness of Inactivated COVID-19 Vaccines Against Symptomatic, Pneumonia, and Severe Disease Caused by the Delta Variant: Real World Study and Evidence - China, 2021. *China CDC Wkly.* **4**, 57–65. (doi:10.46234/ccdcw2022.009)

12. Ye W *et al.* 2023 Inactivated vaccine effectiveness against symptomatic COVID-19 in Fujian, China during the Omicron BA.2 outbreak. *Front. Public Health* **Volume 11-2023**. (doi:10.3389/fpubh.2023.1269194)

13. Li R, Pei S, Chen B, Song Y, Zhang T, Yang W, Shaman J. 2020 Substantial undocumented infection facilitates the rapid dissemination of novel coronavirus (COVID-19). *MedRxiv Prepr. Serv. Health Sci.* , 2020.02.14.20023127. (doi:10.1101/2020.02.14.20023127)

14. Anderson JL. 2001 An ensemble adjustment kalman filter for data assimilation. *Mon. Weather Rev.* **129**, 2884–2903. (doi:10.1175/1520-0493(2001)129<2884:AEAKFF>2.0.CO;2)

15. Pei S, Yamana TK, Kandula S, Galanti M, Shaman J. 2021 Burden and characteristics of COVID-19 in the United States during 2020. *Nature* **598**, 338–341. (doi:10.1038/s41586-021-03914-4)

16. Hao X, Cheng S, Wu D, Wu T, Lin X, Wang C. 2020 Reconstruction of the full transmission dynamics of COVID-19 in Wuhan. *Nature* **584**, 420–424. (doi:10.1038/s41586-020-2554-8)

17. Hale T *et al.* 2021 A global panel database of pandemic policies (Oxford COVID-19 Government Response Tracker). *Nat. Hum. Behav.* **5**, 529–538. (doi:10.1038/s41562-021-01079-8)

18. Guo Z, Chen Y, Xiao G, Jiang H, Yang Z, Li J, Gong L, Jin M, Wang F. 2025 Evaluating the impact of large-scale nucleic acid testing and home quarantine on a novel emerging infectious disease prevention and control: a dynamic modeling approach. *Front. Public Health* **13**. (doi:10.3389/fpubh.2025.1447738)

19. China NHC of the PR of. 2021 Technical Guidelines for COVID-19 Vaccination (Version 1).

20. Agency XN. 2021 Guangzhou: COVID-19 Booster Vaccination Campaign in Progress.

21. Zhao Y *et al.* 2022 Quantifying human mixing patterns in Chinese provinces outside Hubei after the 2020 lockdown was lifted. *BMC Infect. Dis.* **22**, 483. (doi:10.1186/s12879-022-07455-7)

22. Huang Z *et al.* 2023 Effectiveness of inactivated COVID-19 vaccines among older adults in Shanghai: retrospective cohort study. *Nat. Commun.* **14**, 2009. (doi:10.1038/s41467-023-37673-9)

23. Pilz S, Theiler-Schwetz V, Trummer C, Krause R, Ioannidis JPA. 2022 SARS-CoV-2 reinfections: Overview of efficacy and duration of natural and hybrid immunity. *Environ. Res.* **209**, 112911. (doi:10.1016/j.envres.2022.112911)

#### 8. Supplementary Tables

**Table S1. The redefined start date of vaccination dataset.**

| Data in OWD | Definition | First available date | Redefined start date | Note |
|---|---|---|---|---|
| **People vaccinated** | The number of individuals who received at least one dose | 10 June 2021 | 15 December 2020 | Consistent with the start date of total dose data |
| **People fully vaccinated** | The number of individuals who completed the initial vaccination protocol | 12 August 2021 | 19 January 2021 | Since the initial vaccination protocol require individuals to receive two doses of vaccine, with the interval between the two doses being greater than 3 weeks and no more than 8 weeks[19]. I redefined the start date to 19 Jan 2021 (5 weeks after 15 December 2020). |
| **Vaccine booster doses** | The number of individuals who received a booster dose | 5 November 2021 | 19 October 2021 | On 19 October 2021, official news showed that booster doses had been started in some places[20]. |

**Table S2. Parameters of hypergraph-based model**

| Parameter | Description | Prior | Assumed value / Estimates |
|---|---|---|---|
| $\boldsymbol{S_i(t)}$ | Susceptible population in province $i$ at day $t$ | – | – |
| $\boldsymbol{V_i(t)}$ | Vaccinated population in province $i$ at day $t$ | – | – |
| $\boldsymbol{E_i(t)}$ | Exposed population in province $i$ at day $t$ | – | – |
| $\boldsymbol{I_i^r(t)}$ | Reported infectious in province $i$ at day $t$ | – | – |
| $\boldsymbol{I_i^u(t)}$ | Unreported infectious in province $i$ at day $t$ | – | – |
| $\boldsymbol{R_i(t)}$ | Recovered population in province $i$ at day $t$ | – | – |
| $\boldsymbol{N_i(t)}$ | Total population in province $i$ at day $t$ | – | – |
| $\boldsymbol{\Theta_i}(\cdot)$ | Force of infection in province $i$ | – | – |
| $\boldsymbol{C_i(t)}$ | Nonlinear infection proportionality coefficient at day $t$ in province $i$ | – | – |
| $\boldsymbol{A_i(t)}$ | Baseline of $C_i(t)$ | (0.05,0.13) (1 January 2020 – 11 April 2021)<br>(0.05,0.30) (19 July 2021 – 21 November 2021)<br>(0.05,0.48) (28 February 2022 – 10 November 2022) | Province-specific initial distributions were obtained from the posterior ensemble of the EAKF after assimilating the first 50 days of data. |
| $\boldsymbol{v_i}$ | Nonlinear infection kernel in province $i$ | (0.8,2) | Province-specific initial distributions were obtained from the posterior ensemble of the EAKF after assimilating the first 50 days of data. |
| $\boldsymbol{d}$ | Average number of contact clusters per day for each individual | – | 1.19 [21] |
| $\boldsymbol{P_i(t)}$ | Stringency index at day $t$ in province $i$ | – | – |
| $\boldsymbol{M_{ij}(t)}$ | Number of human mobility at day $t$ from province $j$ to province $i$ | – | – |
| $\boldsymbol{\eta_i(t)}$ | Number of fully vaccinations dose at day $t$ in province $i$ | – | – |
| $\boldsymbol{b_i(t)}$ | Number of booster vaccinations at day $t$ in province $i$ | – | – |
| $\boldsymbol{\varepsilon(t)}$ | Vaccination effectiveness at day $t$ | – | 0.79 (15 December 2020 – 11 April 2021) [10] |

| | | | |
|---|---|---|---|
| | | | 0.51 (19 July 2021 – 21 November 2021) [11]<br>0.27 (28 February 2022 – 10 November 2022) [12] |
| $\boldsymbol{\lambda}$ | Vaccine-induced immunity waning rate | – | 1/180 days [22] |
| $\boldsymbol{\gamma}$ | Infection-induced immunity waning rate | – | 1/365 days [23] |
| $\boldsymbol{\alpha_i(t)}$ | Ascertainment rate at day $t$ in province $i$ | (0,1) | 0.14 (0.11 − 0.17) [16] |
| $\boldsymbol{Z}$ | Average latency period | (2,5) | 2.94 (2.05 − 4.36) |
| $\boldsymbol{D}$ | Average duration of infection | (5,10) | 5.54 (5.03 − 6.73) |

**Table S3: Sensitivity analysis of model inference to the mobility multiplicative factor ($\delta$) and average degree ($d$)**

| Scenario | Baseline | $\delta_1 = 1.1\delta$ | $\delta_2 = 0.9\delta$ | Average degree $d = 2$ |
|---|---|---|---|---|
| **Mobility multiplicative factor ($\delta$)** | $3.09 \times 10^5$ | $3.39 \times 10^5$ | $2.78 \times 10^5$ | $3.09 \times 10^5$ |
| **Average cluster number ($d$)** | 1.19 | 1.19 | 1.19 | 2 |
| **MAE (national)** | 563.20 | 536.30 | 537.61 | 559.20 |
| **Gini (1 January 2020 – 31 March 2020)** | 0.29 | 0.29 | 0.29 | 0.29 |
| **Gini (1 April 2020 – 11 April 2021)** | 0.25 | 0.23 | 0.25 | 0.25 |
| **Gini (12 April 2021 – 21 November 2021)** | 0.32 | 0.32 | 0.32 | 0.32 |
| **Gini (22 November 2021 – 10 November 2022)** | 0.30 | 0.30 | 0.30 | 0.32 |
| **Peak of $R_e$ (1 January 2020 – 31 March 2020)** | 1.70(95%CrI 1.42 - 1.98) | 1.70(95%CrI 1.42 - 1.98) | 1.70(95%CrI 1.42 - 1.98) | 2.85(95%CrI 2.39 - 3.33) |
| **Peak of $R_e$ (1 April 2020 – 11 April 2021)** | 0.93(95%CrI 0.78 - 1.08) | 0.95(95%CrI 0.79 - 1.09) | 0.94(95%CrI 0.78 - 1.08) | 1.53(95%CrI 1.27 - 1.76) |
| **Peak of $R_e$ (12 April 2021 – 21 November 2021)** | 1.00(95%CrI 0.80 - 1.17) | 1.01(95%CrI 0.84 - 1.13) | 1.00(95%CrI 0.80 - 1.19) | 1.55(95%CrI 1.24 - 1.85) |
| **Peak of $R_e$ (22 November 2021 – 10 November 2022)** | 1.84(95%CrI 1.49 - 2.13) | 1.92(95%CrI 1.56 - 2.21) | 1.72(95%CrI 1.37 - 1.98) | 2.80(95%CrI 2.25 - 3.26) |

#### 9. Supplementary Figures

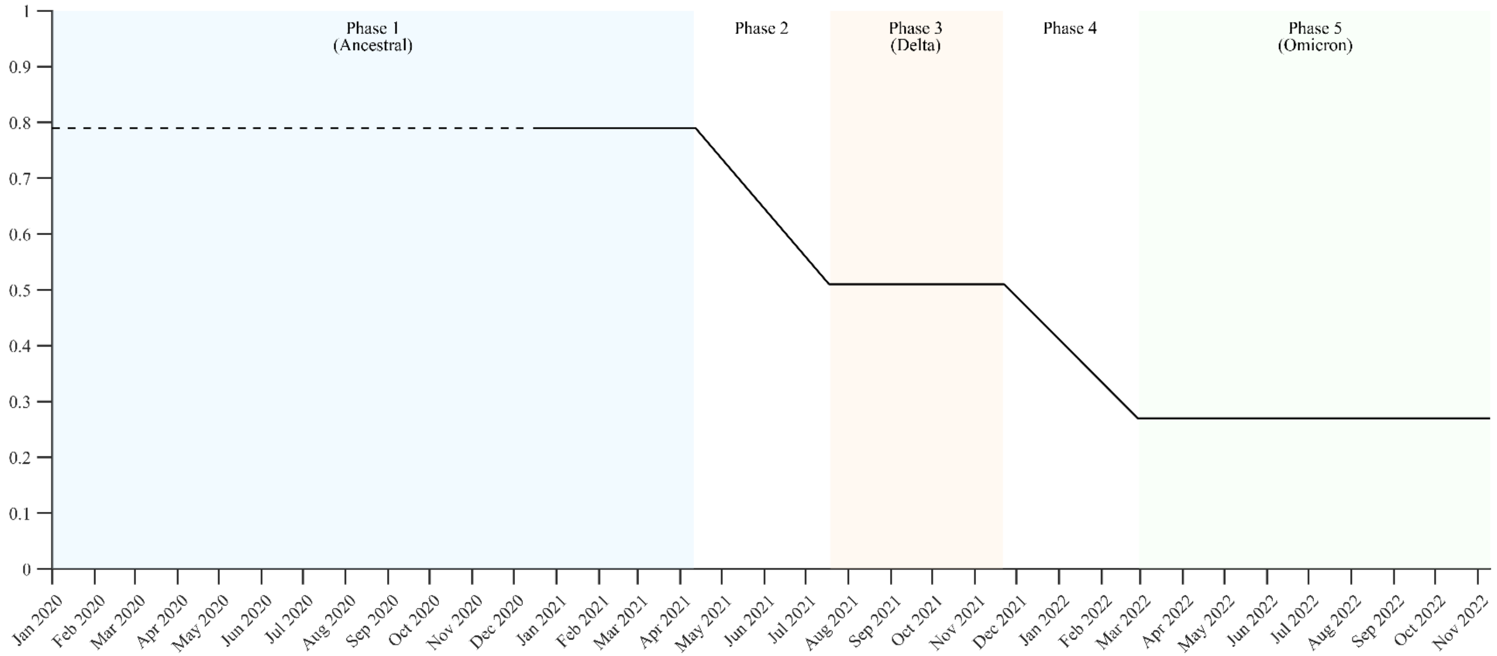

**Figure S1**. Time-varying vaccination effectiveness $\varepsilon(t)$. The black line is shown as a horizontal dashed line before 15 December 2020, indicating that $\varepsilon(t)$ is not applicable prior to vaccine rollout; from 15 December 2020 onward, it is shown as a solid line.

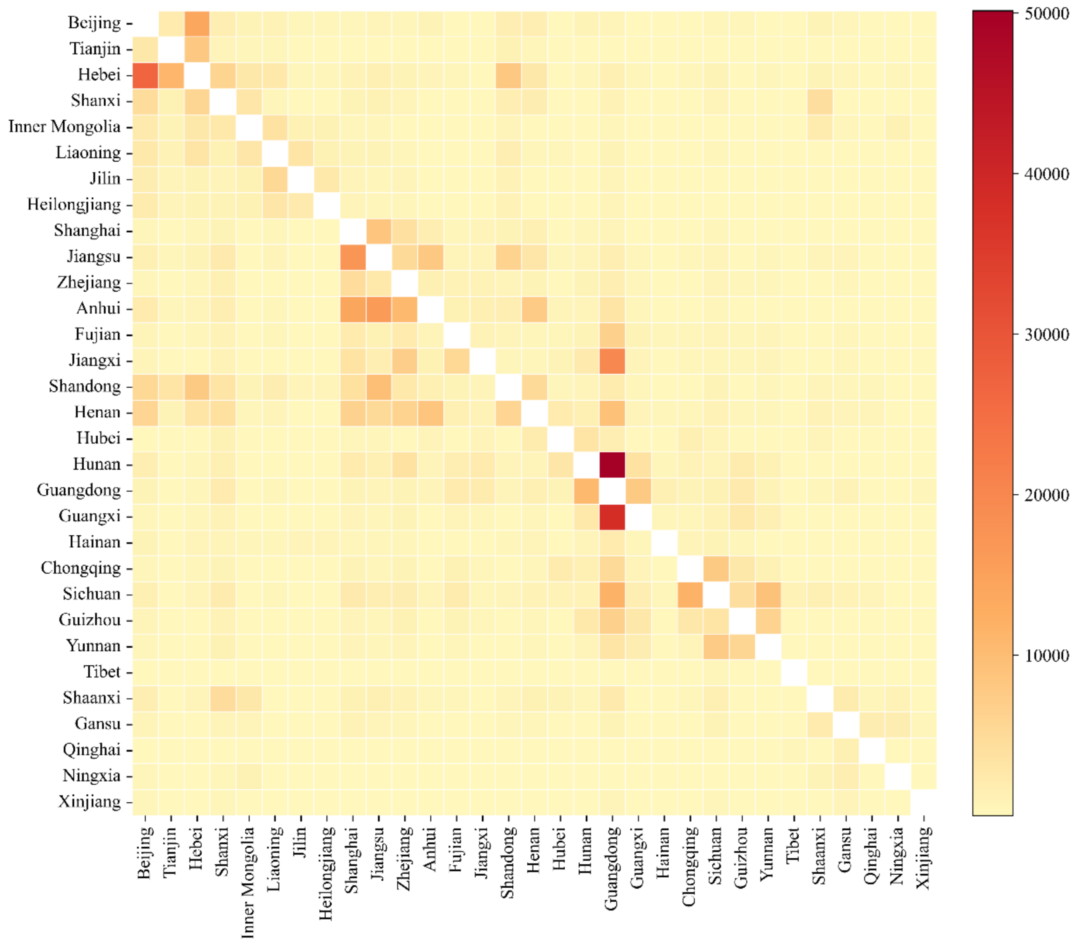

**Figure S2**. Inter-provincial mobility flows in mainland China on 13 February 2020, the peak of the first epidemic wave. Provinces are ordered according to the standard provincial administrative division code sequence of mainland China, which is widely used in official statistics and national data reporting.

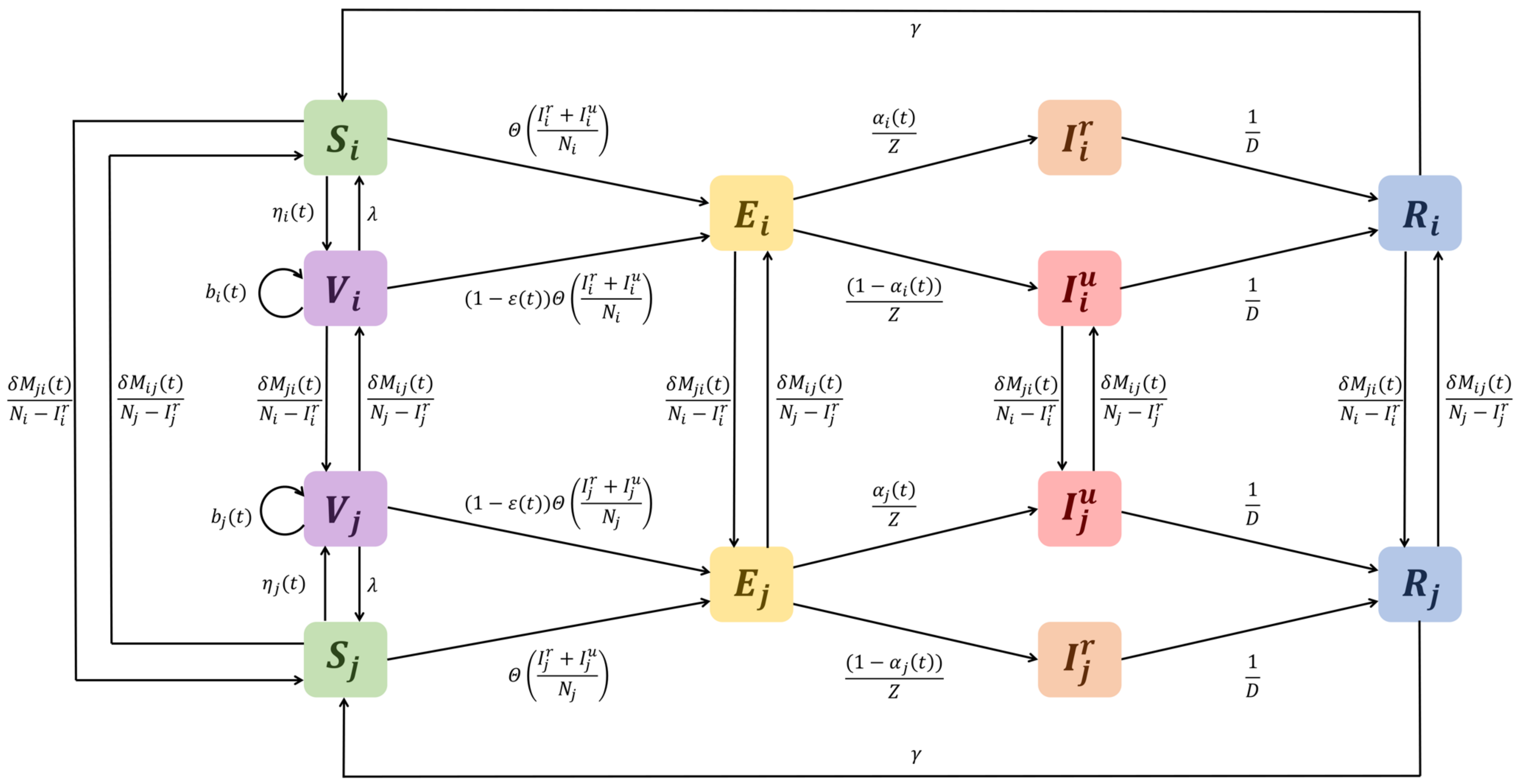

**Figure S3**. Schematic of SVEIRS Compartmental model.

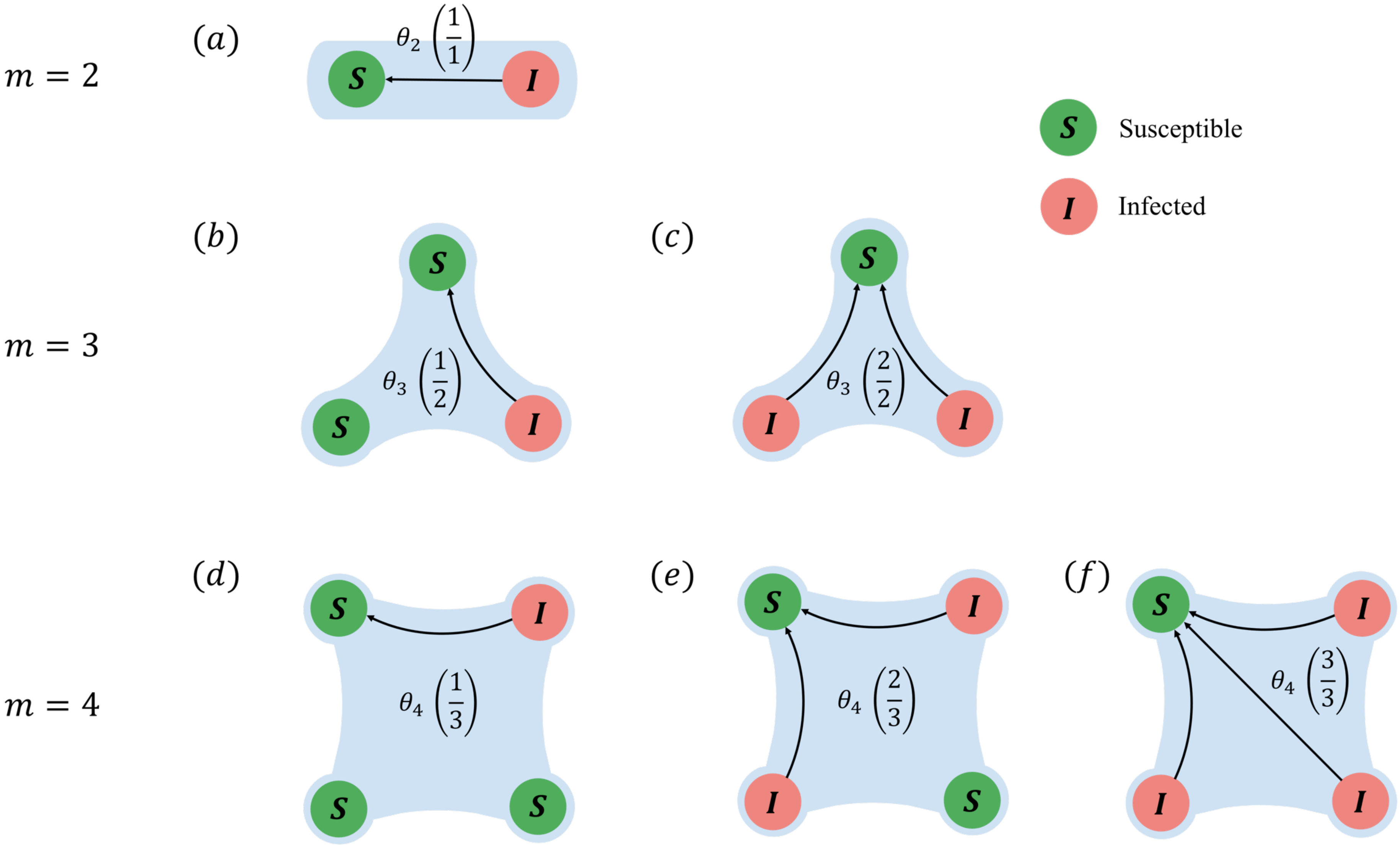

**Figure S4**. Examples of the force of infection to a susceptible in the *m*-size cluster.

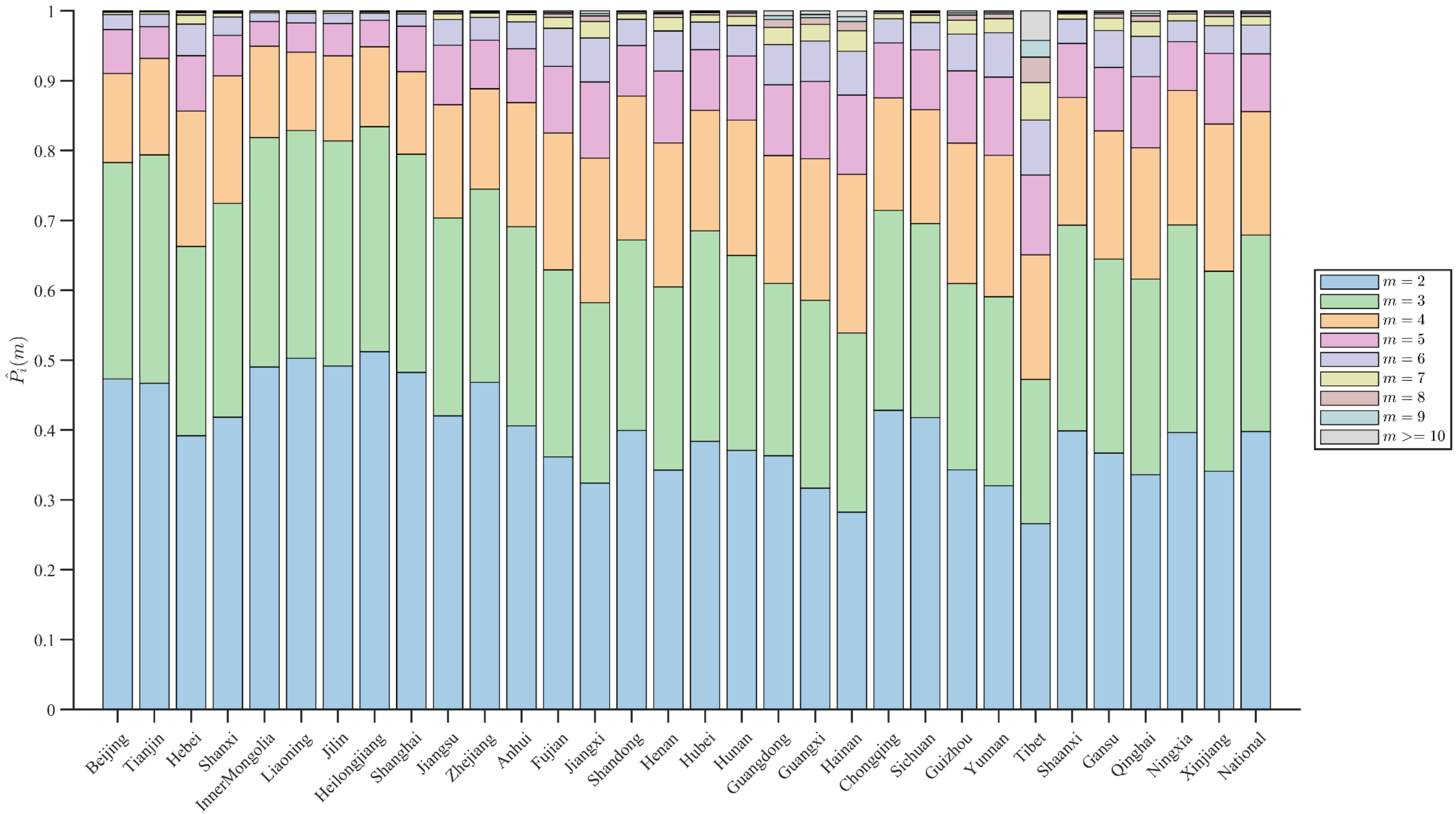

**Figure S5**. Household scale distribution $\hat{P}_i(m)$ in mainland China.

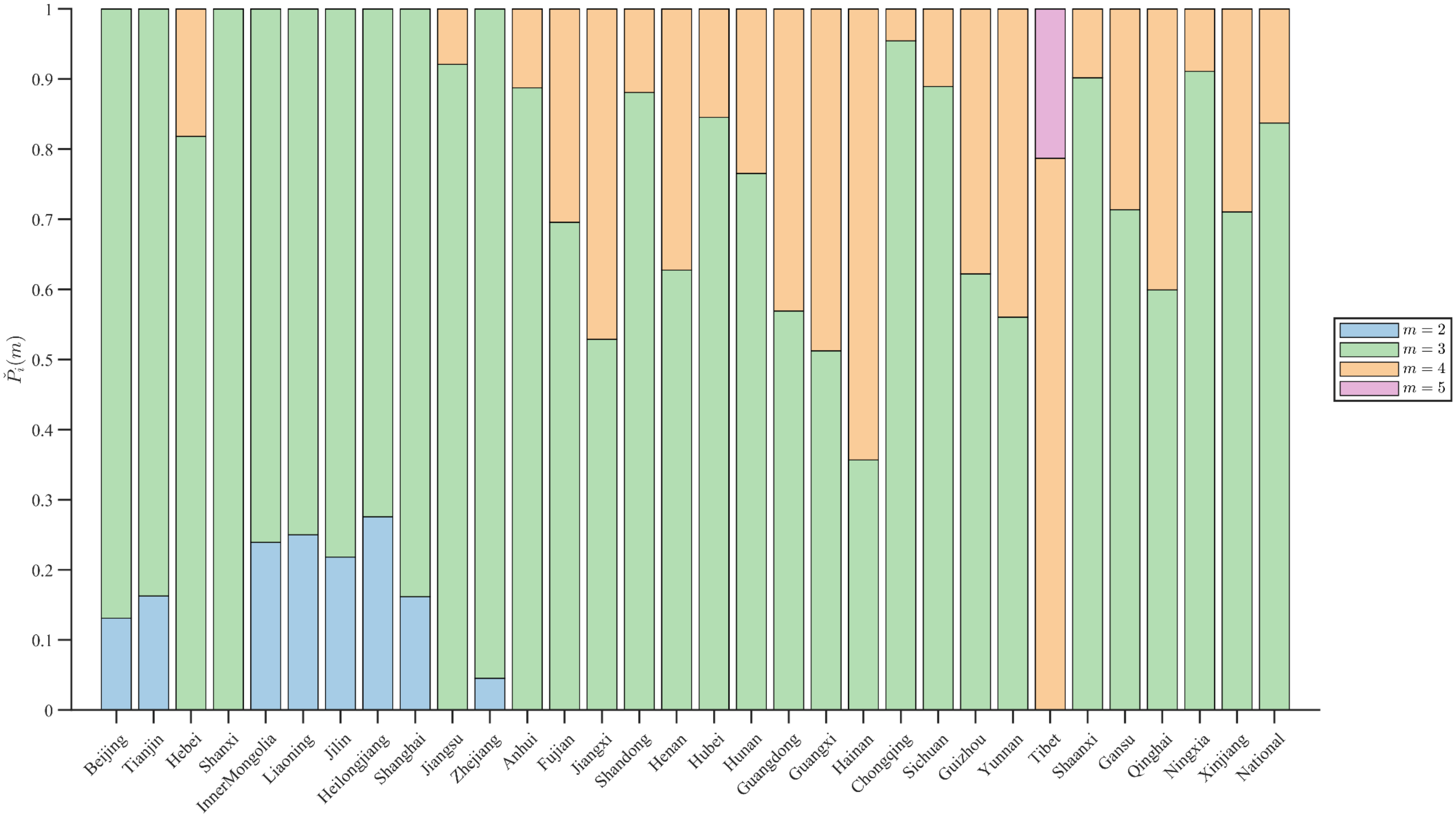

**Figure S6**. Modified minimized-variance cluster size distribution $\check{P}_i(m)$ in mainland China.

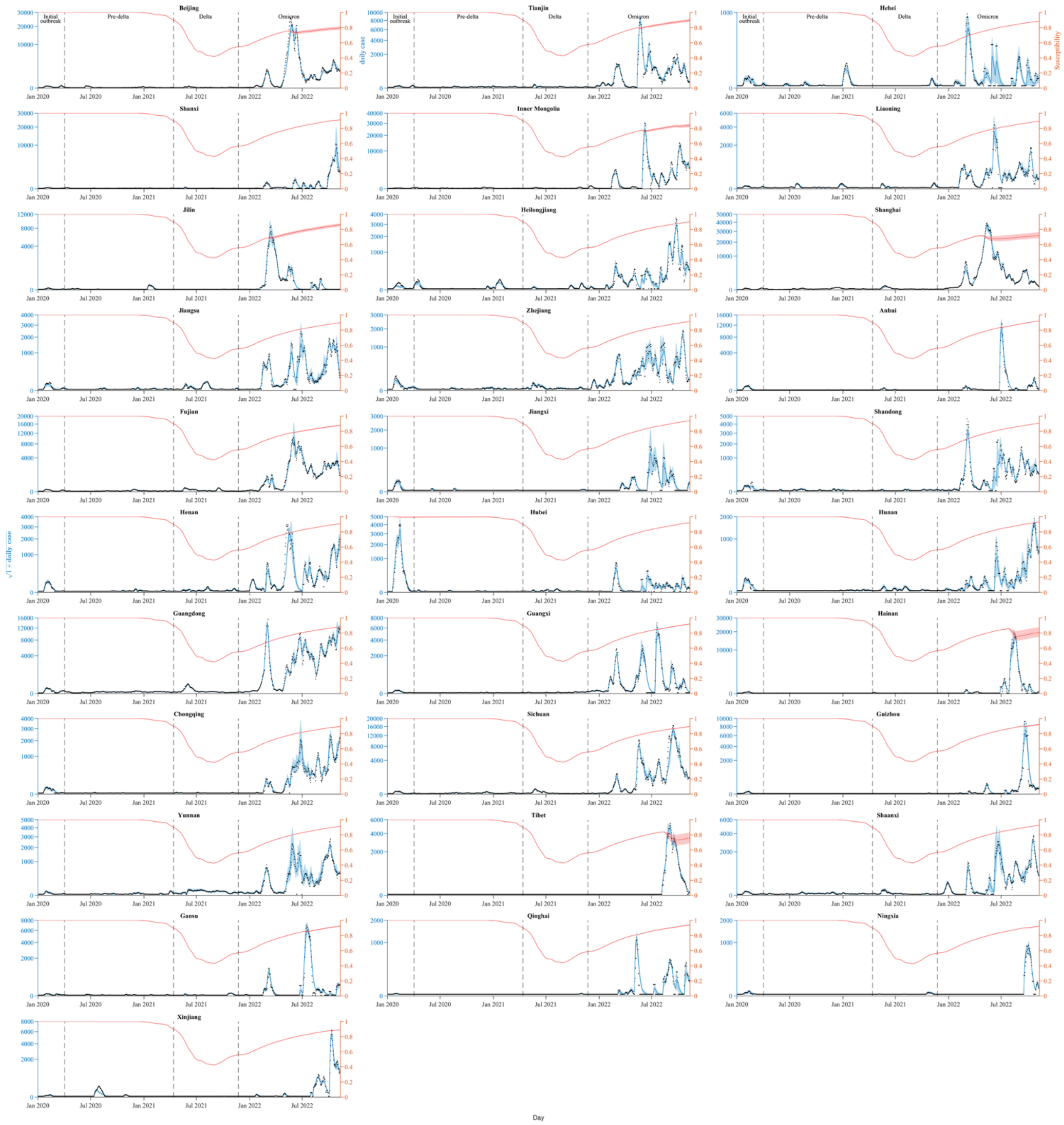

**Figure S7**: Model fitting to daily case numbers (black dots) and susceptibility (red lines) in 31 provinces of mainland China. The blue shades represent the 95%CrI of estimated daily reported cases. The solid red lines and red shades show the median and 95%CrI of estimated susceptibility.

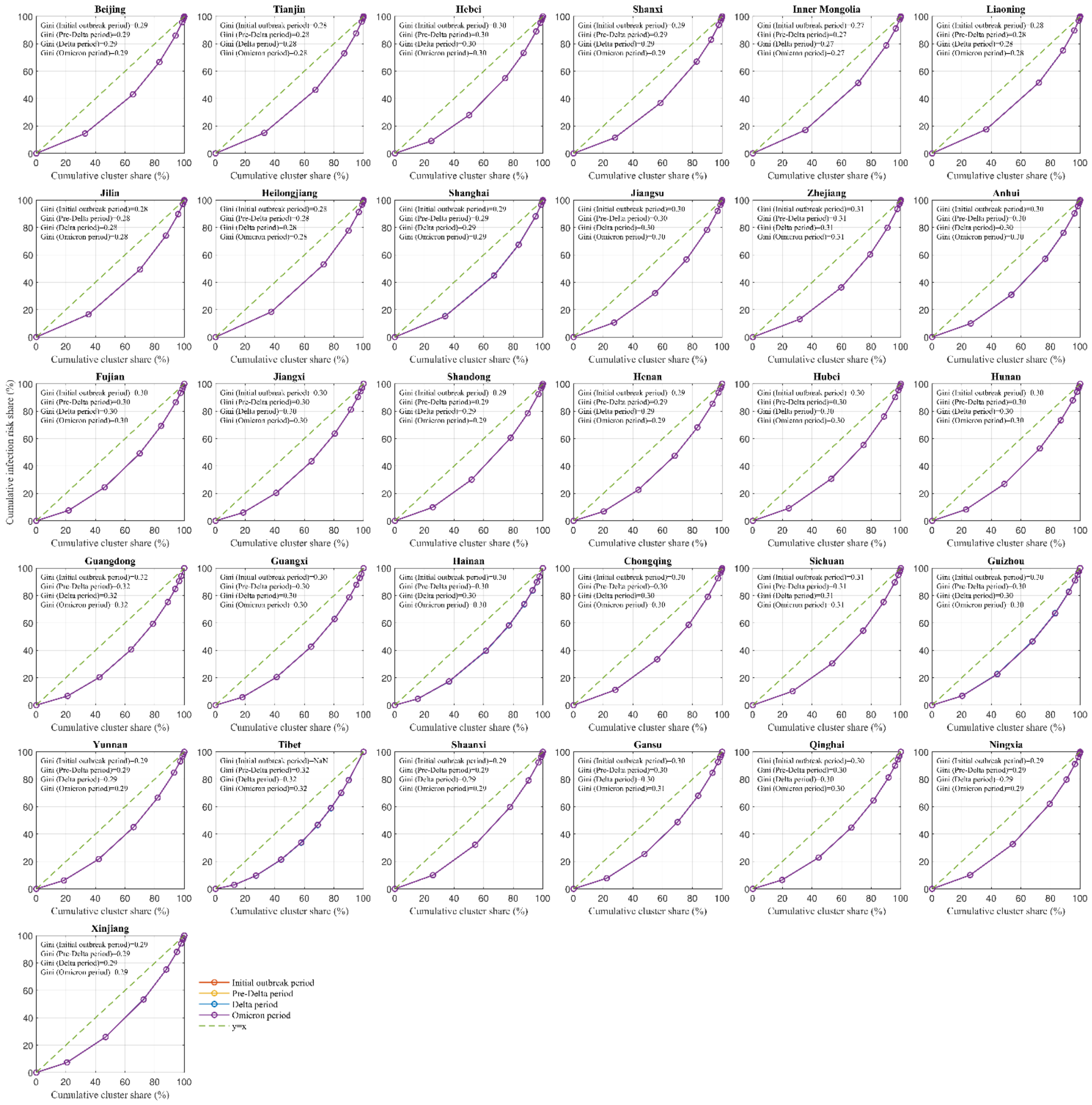

**Figure S8**: Lorenz curves depict the cumulative distribution of infection risk across cluster sizes for four study periods in province-level of mainland China. The diagonal line denotes perfect equality. Gini coefficients summarize the degree of risk heterogeneity in each period.

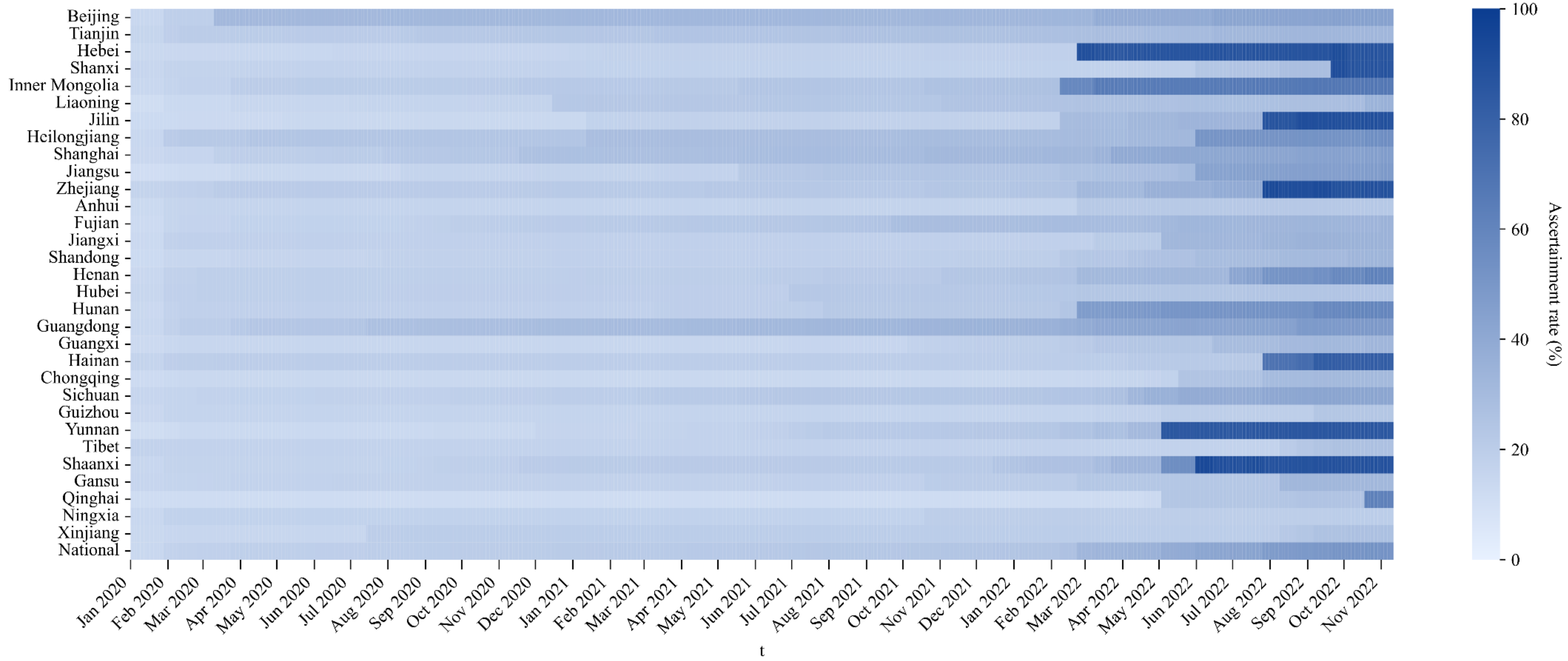

**Figure S9**: Spatiotemporal evolution of the ascertainment rate in the provinces of mainland China from 1 January 2020 to 10 November 2022. The national ascertainment rate is obtained by taking the population-weighted average of provincial ascertainment rates.

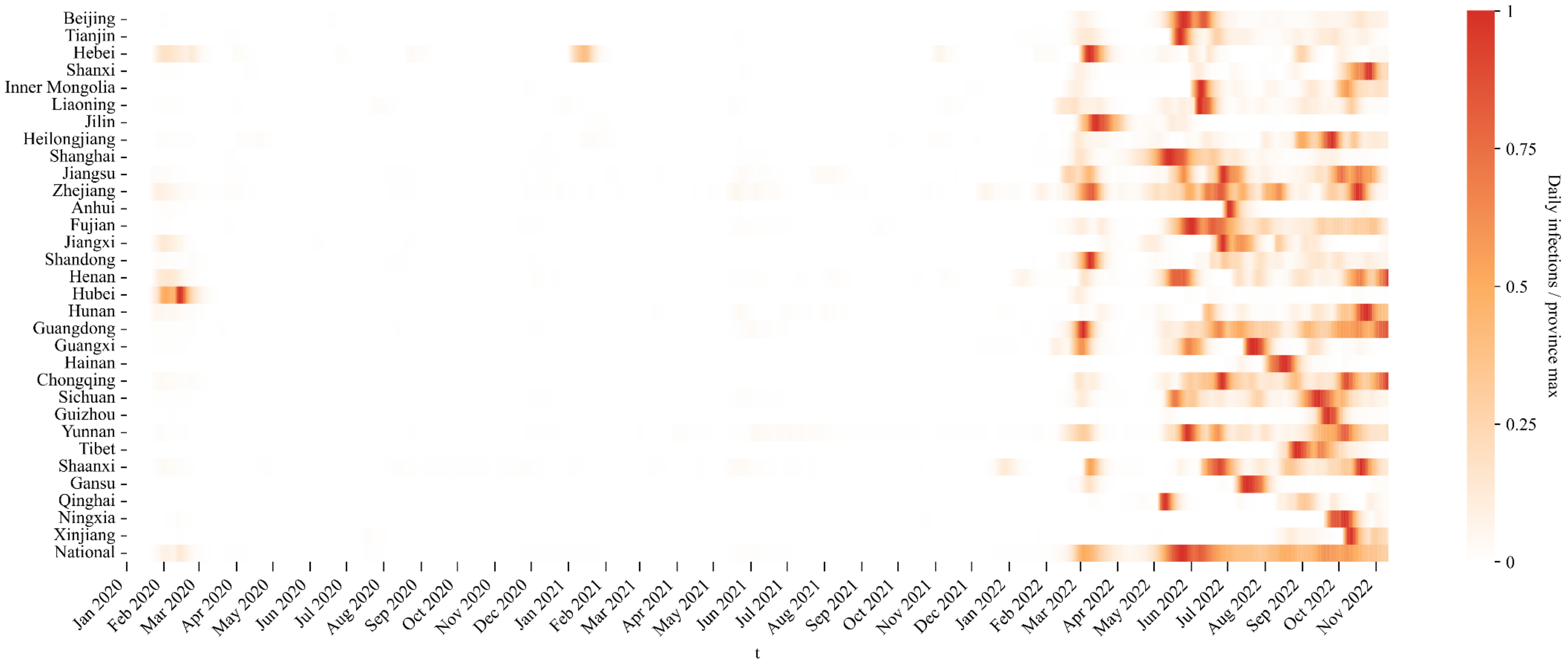

**Figure S10**. Normalized estimated daily infections at provinces of mainland China from 1 January 2020 to 10 November 2022. For each province, daily new infections were scaled to [0,1] by dividing by the province-specific maximum over the study period. The national value is obtained by normalizing the sum of provincial estimated daily infections.

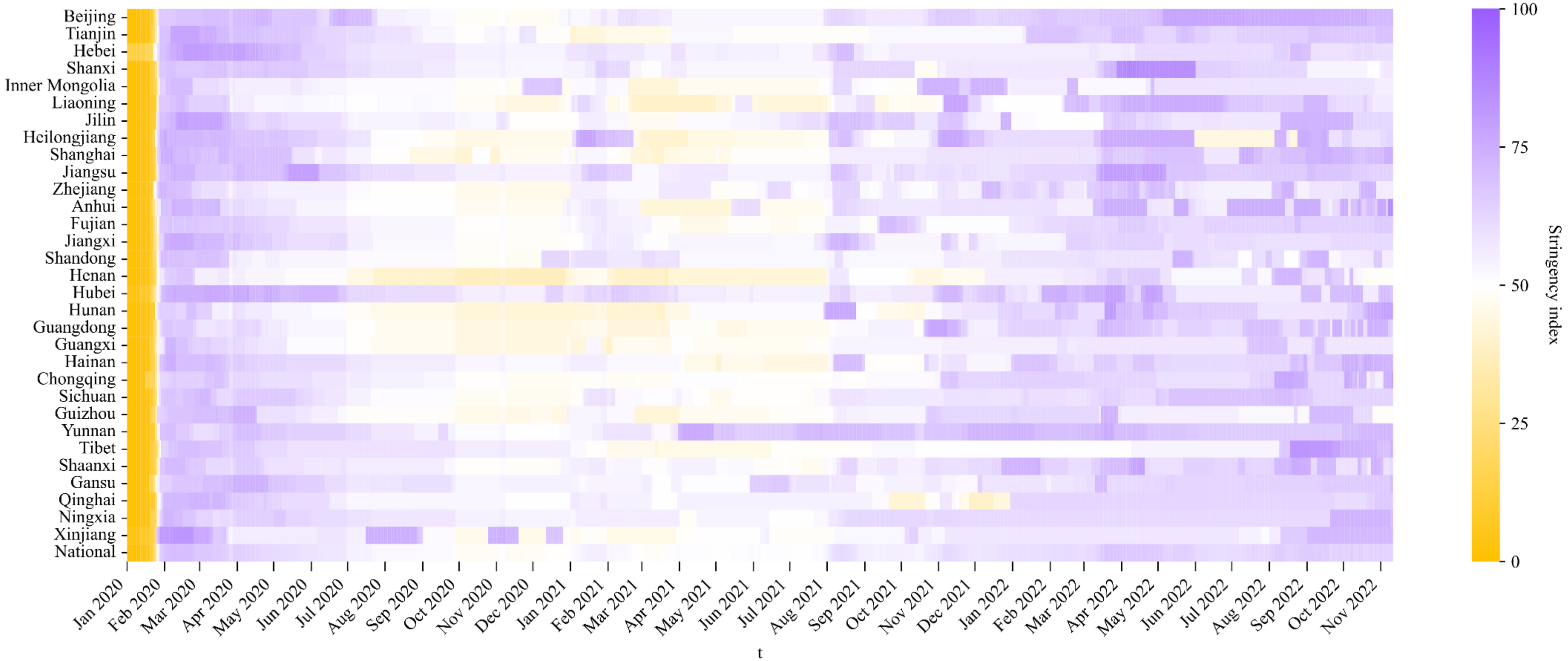

**Figure S11**: Stringency index at provinces of mainland China from 1 January 2020 to 10 November 2022. The national stringency index is obtained by taking the population-weighted average of provincial stringency index.

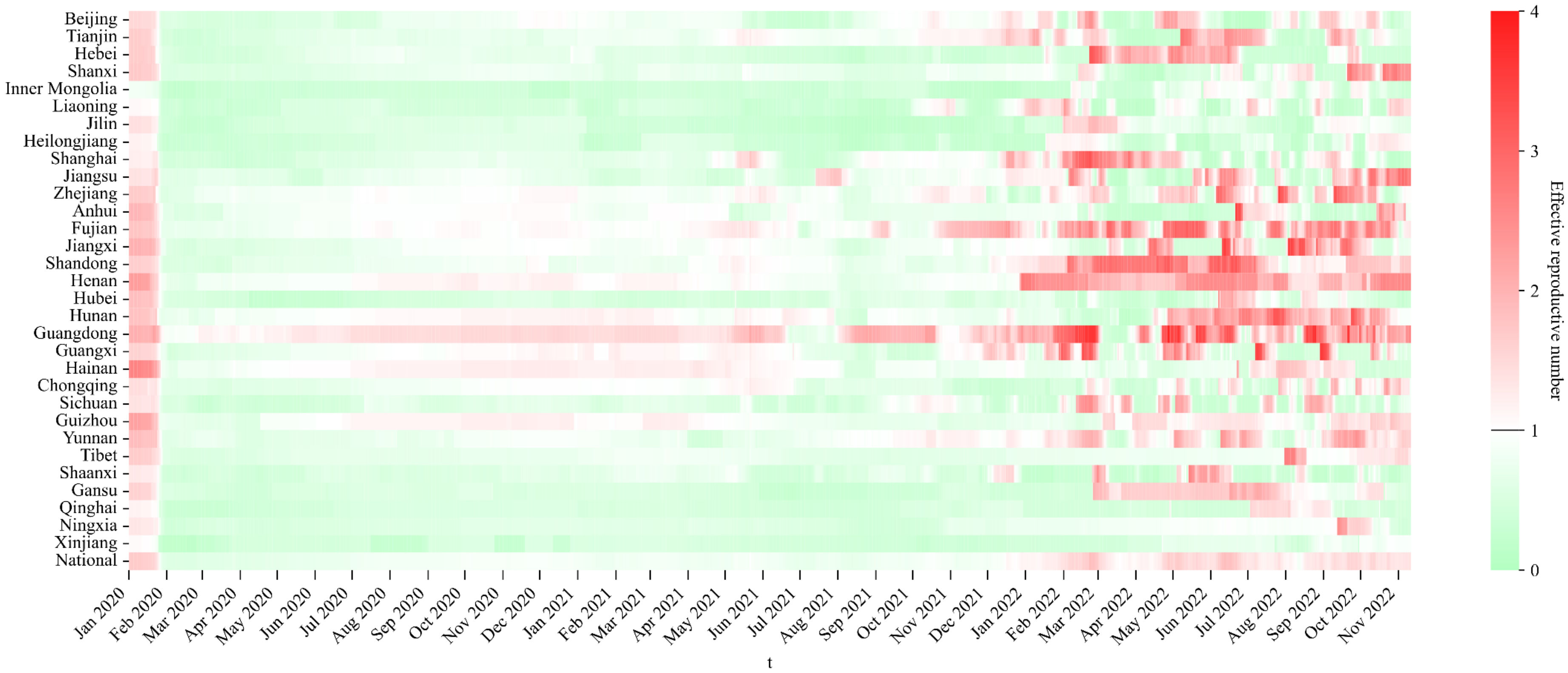

**Figure S12**: Spatiotemporal evolution of effective reproductive number in the provinces of mainland China from 1 January 2020 to 10 November 2022.

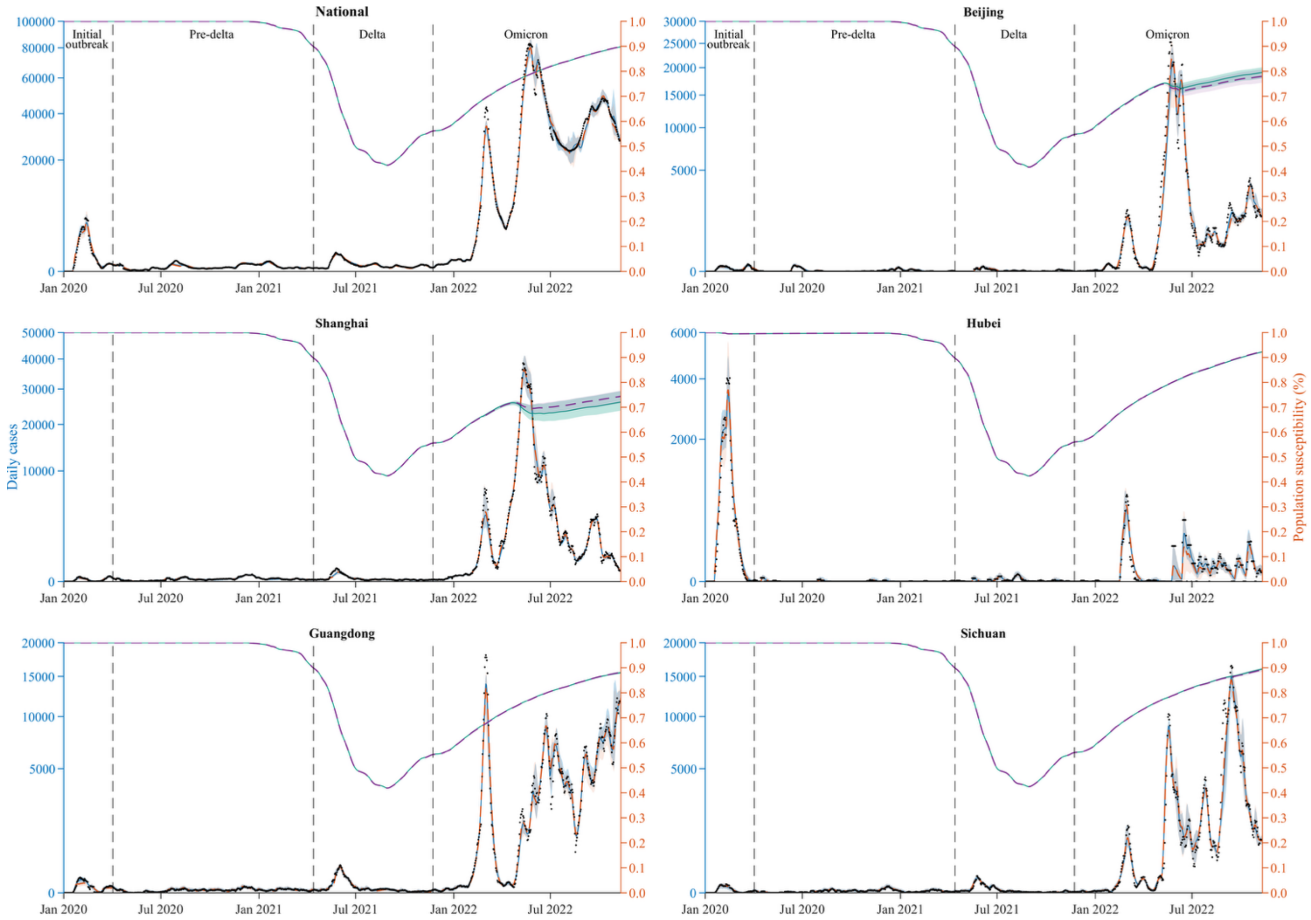

**Figure S13**: Sensitivity analyses results using the mobility multiplicative factor $\delta_1 = 1.1\delta$. Model fitting to daily reported case numbers (black dots) and population susceptibility in mainland China and five provinces (Beijing, Shanghai, Hubei, Guangdong, and Sichuan) from 1 January 2020 to 10 November 2022. These provinces were selected for their epidemiological and demographic representativeness: Beijing as the national capital, Shanghai for its extremely high population density, Hubei as the initial epicenter of the COVID-19 outbreak, Guangdong as the most populous province, and Sichuan as a major transportation hub in western China. Blue (baseline) and orange ($\delta_1 = 1.1\delta$) curves represent the median estimated daily cases, with corresponding shaded areas showing the 95% CrI. Teal (baseline) and purple (alternative inference) curves denote the median estimated susceptibility, with shaded bands indicating the 95% CrI. Solid lines indicate the baseline inference, while dashed lines indicate the alternative inference. Vertical gray dashed lines mark the beginning of the pre-Delta, Delta, and Omicron periods.

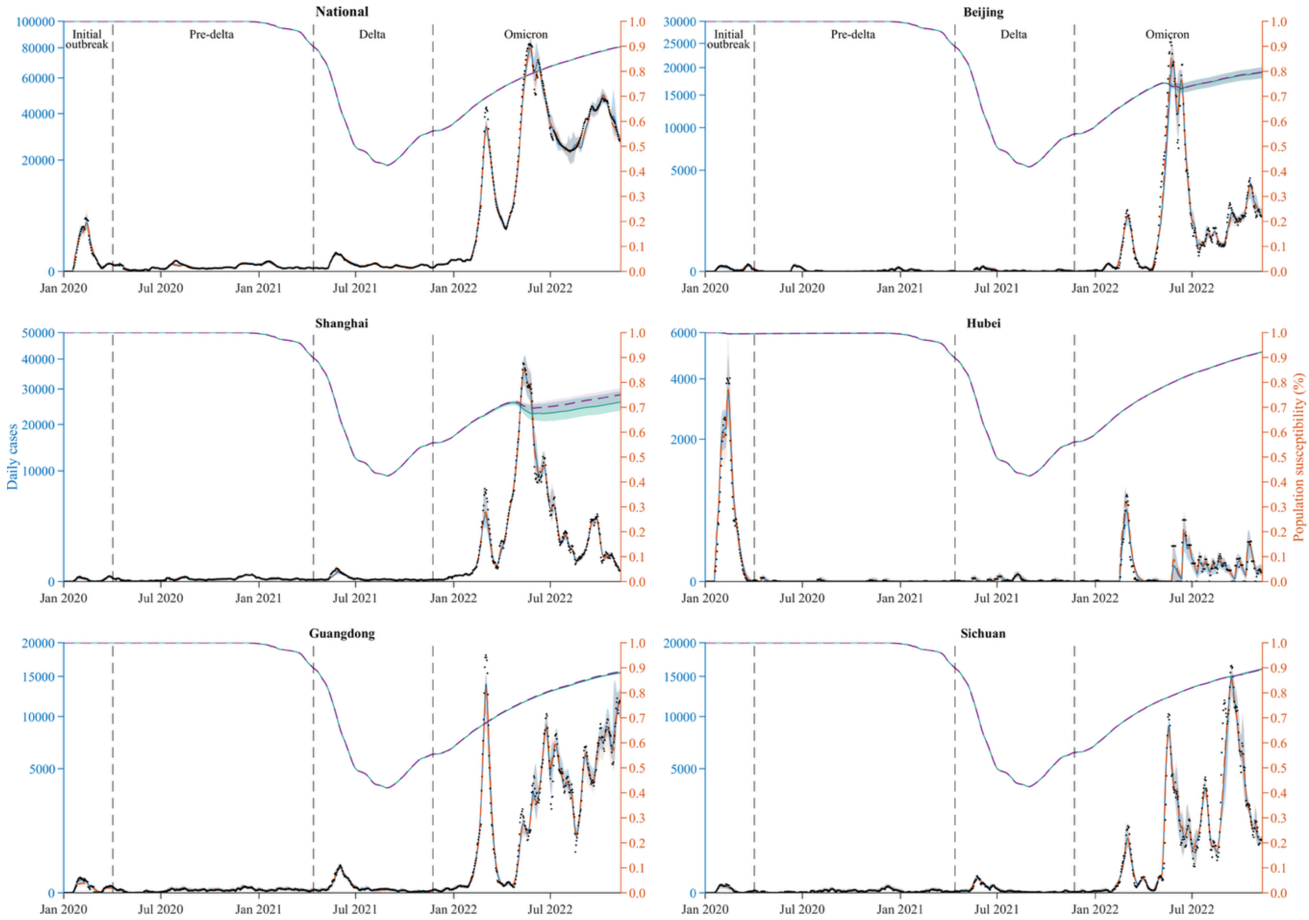

**Figure S14**: Sensitivity analyses results using the mobility multiplicative factor $\delta_2 = 0.9\delta$. Model fitting to daily reported case numbers (black dots) and population susceptibility in mainland China and five provinces (Beijing, Shanghai, Hubei, Guangdong, and Sichuan) from 1 January 2020 to 10 November 2022. These provinces were selected for their epidemiological and demographic representativeness: Beijing as the national capital, Shanghai for its extremely high population density, Hubei as the initial epicenter of the COVID-19 outbreak, Guangdong as the most populous province, and Sichuan as a major transportation hub in western China. Blue (baseline) and orange ($\delta_2 = 0.9\delta$) curves represent the median estimated daily cases, with corresponding shaded areas showing the 95% CrI. Teal (baseline) and purple (alternative inference) curves denote the median estimated susceptibility, with shaded bands indicating the 95% CrI. Solid lines indicate the baseline inference, while dashed lines indicate the alternative inference. Vertical gray dashed lines mark the beginning of the pre-Delta, Delta, and Omicron periods.

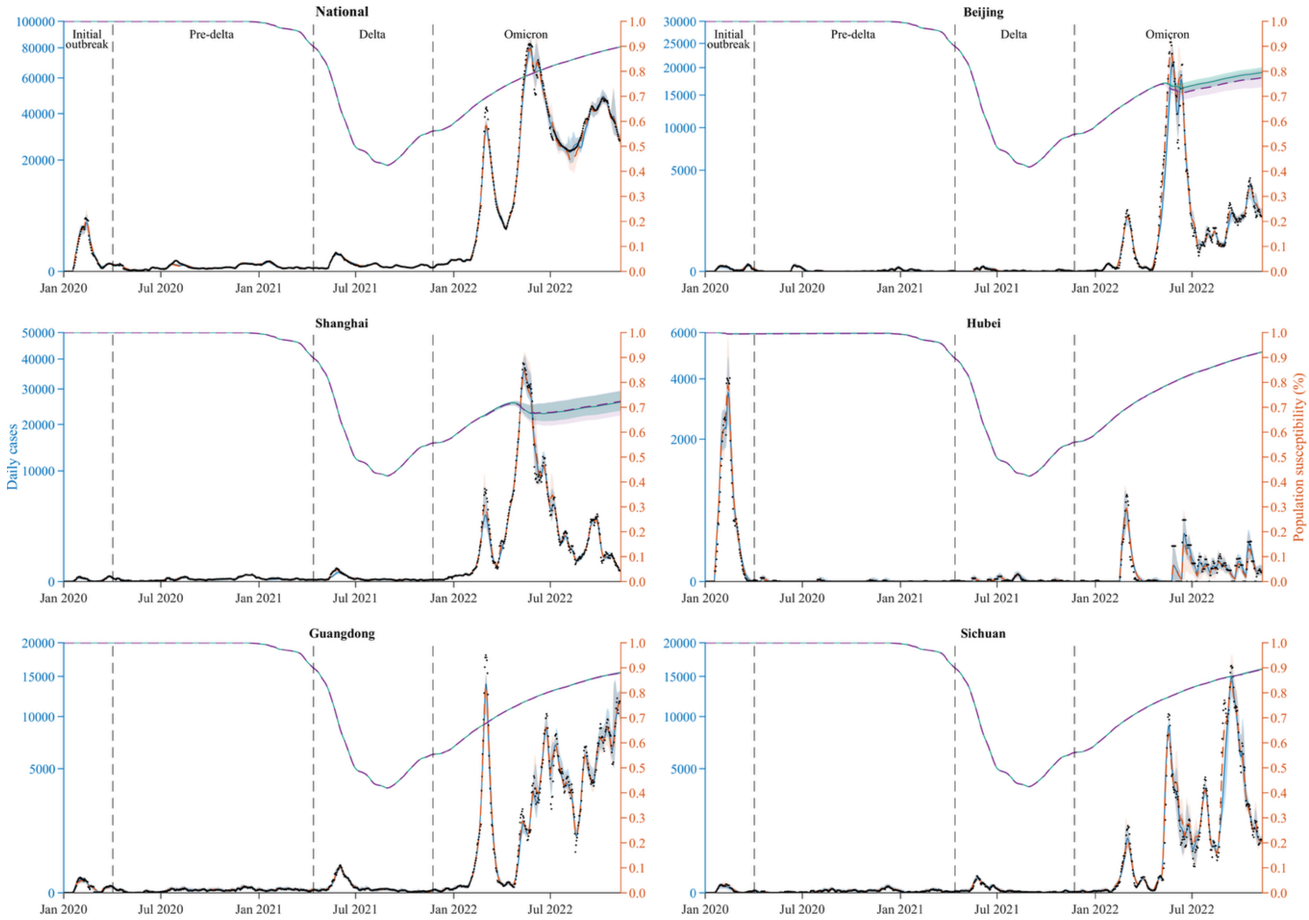

**Figure S15**: Sensitivity analyses results using average degree $d = 2$. Model fitting to daily reported case numbers (black dots) and population susceptibility in mainland China and five provinces (Beijing, Shanghai, Hubei, Guangdong, and Sichuan) from 1 January 2020 to 10 November 2022. These provinces were selected for their epidemiological and demographic representativeness: Beijing as the national capital, Shanghai for its extremely high population density, Hubei as the initial epicenter of the COVID-19 outbreak, Guangdong as the most populous province, and Sichuan as a major transportation hub in western China. Blue (baseline) and orange ($d = 2$) curves represent the median estimated daily cases, with corresponding shaded areas showing the 95% CrI. Teal (baseline) and purple (alternative inference) curves denote the median estimated susceptibility, with shaded bands indicating the 95% CrI. Solid lines indicate the baseline inference, while dashed lines indicate the alternative inference. Vertical gray dashed lines mark the beginning of the pre-Delta, Delta, and Omicron periods.
